# Genetic Heterogeneity Across Dimensions of Alcohol Use Behaviors

**DOI:** 10.1101/2023.12.26.23300537

**Authors:** Jeanne E Savage, Peter B Barr, Tanya Phung, Younga H. Lee, Yingzhe Zhang, COGA Investigators, Tian Ge, Jordan W. Smoller, Lea K. Davis, Jacquelyn Meyers, Bernice Porjesz, Danielle Posthuma, Travis T Mallard, Sandra Sanchez-Roige

## Abstract

**Background:** Increasingly large samples in genome-wide association studies (GWAS) for alcohol use behaviors (AUBs) have led to an influx of implicated genes, yet the clinical and functional understanding of these associations remains low. This is, in part, because most GWASs do not account for complex and varied manifestations of AUBs. This study applied a multidimensional framework to investigate the latent genetic structure underlying heterogeneous dimensions of AUBs.

**Methods:** Multi-modal assessments (self-report, interview, electronic health records) were obtained from approximately 400,000 UK Biobank participants. GWAS was conducted for 18 distinct AUBs, including consumption, drinking patterns, alcohol problems, and clinical sequelae. Latent genetic factors were identified and carried forward to GWAS using genomic structural equation modeling, followed by functional annotation, genetic correlation, and enrichment analyses to interpret the genetic associations.

**Results:** Four latent factors were identified: *Problems, Consumption, BeerPref* (declining alcohol consumption with a preference for drinking beer), and *AtypicalPref* (drinking fortified wine and spirits). The latent factors were moderately correlated (*r_g_=*.12-.57) and had distinct patterns of associations, with *BeerPref* in particular implicating many novel genomic regions. Patterns of regional and cell type specific gene expression in the brain also differed between the latent factors.

**Conclusion:** Deep phenotyping and multi-modal assessment is an important next step to improve understanding of the genetic etiology of AUBs, in addition to increasing sample size. Further effort is required to uncover the genetic heterogeneity underlying AUBs using methods that account for their complex, multidimensional nature.

## Introduction

Alcohol use behaviors (AUBs) encompass a broad spectrum of normative and problematic activities related to the consumption of alcohol which, collectively, have an enormous impact on human health and well-being (World Health Organization, 2018). While drinking is a clear prerequisite for developing alcohol-related problems, there are important distinctions between different dimensions of AUBs, such as quantity and patterns of consumption, acute episodic (binge) drinking, and problematic alcohol use (PAU). PAU itself describes a variety of risky or maladaptive drinking behaviors that may merit a clinical diagnosis of alcohol use disorder (AUD).

Although AUBs have varying clinical and epidemiological correlates (Gunn, Finn, Endres, Gerst, & Spinola, 2013; Savage & Dick, 2023; Smith, Shevlin, Murphy, & Houston, 2010), all of its diverse forms have a substantial heritable component, with twin studies suggesting that genetics account for approximately 40-60% of individual differences (Dick, Meyers, Rose, Kaprio, & Kendler, 2011; Verhulst, Neale, & Kendler, 2015). Large-scale genome-wide association studies (GWASs) have begun to yield success in identifying the specific genes underlying the heritability of AUBs (Deak & Johnson, 2021), particularly for normative drinking (Liu et al., 2019; Mallard et al., 2022) and PAU (Zhou et al., 2023; Zhou et al., 2020).

In recent years, the foremost strategy for GWAS has centered on increasing sample sizes to boost statistical power, aiming to detect genetic variants with subtle effects. This perspective has often prompted researchers to prioritize studying phenotypes that have been measured in large samples, even if the available phenotypic measures are shallow or unidimensional. This strategy has proven successful at increasing the number of variants and genomic loci associated with AUBs (Gelernter et al., 2019; Kranzler et al., 2019; Liu et al., 2019; Sanchez-Roige et al., 2019; Walters et al., 2018; Zhou et al., 2023; Zhou et al., 2020), but it has proven challenging to advance clinical or functional understanding of these statistical associations. For example, in comparison with the single significant locus found in a GWAS of alcohol dependence (AD) in approximately 50,000 individuals (Walters et al., 2018), a GWAS of PAU in over 400,000 individuals (Zhou et al., 2020) increased the number of associated risk loci to 29. However, the increase in accuracy of out-of-sample prediction of AD/AUD was only modest (0.5% - 1.7% from Walters et al. versus 0.8% - 2% from Zhou et al.). Further increasing the PAU sample size to over 1 million individuals resulted in 110 associated loci (Zhou et al., 2023), but this gain did not translate into improved predictive accuracy (0.15%) or immediately actionable biological insights.

These findings point to diminishing returns on ever-larger sample size investments for the unidimensional AUBs typically studied with GWAS (i.e., binary AUD diagnostic status or overall consumption quantity). While the reasoning for focusing on such phenotypes can be understood through the lens of increasing statistical power, numerous twin and molecular studies have provided evidence for a distinct genetic architecture both within and between dimensions of AUBs (Dick et al., 2011; Kendler, Aggen, Prescott, Crabbe, & Neale, 2012; Sanchez-Roige et al., 2019). It is plausible that when genetically heterogeneous measures are combined together, the association of individual variants becomes diluted, leading to smaller and more uncertain effect sizes (which can be identified with sufficient statistical power) and less accurate individual-level prediction. Approaches to account for heterogeneity, such as item-level genetic analysis, have demonstrated that gene identification and interpretation can be improved by sharpening the resolution of the AUB outcomes under investigation (Mallard et al., 2022). However, few well-powered studies have applied such approaches to the diverse spectrum of AUBs, particularly for measures of alcohol-related problems that are of most clinical relevance. In the current study, we carried out a multimodal, multidimensional analysis of a broad set of AUBs derived from a large biobank sample, most of which have not been targeted in prior GWAS investigations. We used structural equation modeling and bioinformatic tools to carve out the boundaries of the genetic architecture underlying numerous AUBs and to characterize the shared and unique genetic influences across these dimensions.

## Methods

### Sample

Data was drawn from the UK Biobank (UKB) (Bycroft et al., 2018), a population-based sample of approximately 500,000 adults from the UK with self-report surveys, linked electronic health records from the national health registry databases (accessed April 19, 2022), and genotypic data from imputed genome-wide microarrays. The National Research Ethics Service Committee North West–Haydock ethically approved this initiative (reference 11/NW/0382) and participants provided informed written consent. Data were accessed under application #16406.

### Univariate GWAS of AUBs

Catalogs of all available data were searched manually to identify fields related to alcohol use (e.g., consumption habits), problems (e.g., AUD diagnoses), and alcohol-related sequelae (e.g., cirrhosis). A total of 36 phenotypes was obtained after extracting the data and combining clinical (ICD/READ) diagnostic codes for similar domains, as summarized in **Table S1**. Univariate GWAS was conducted on each phenotype in up to 386,971 unrelated individuals of European (EUR) ancestry (**Table 1**), using either linear or logistic regression in PLINK v2.00 (Chang et al., 2015). Self-reported biological sex, age, genotyping array, and 20 within-ancestry principal components were included as covariates. Full details of the genotyping, quality control, and analysis pipeline have been described previously (Savage et al., 2018). We used linkage disequilibrium score regression (LDSCv1.0.1) (Bulik-Sullivan et al., 2015) to estimate heritability (*h*^2^_SNP_) and calculate genetic correlations (*r_g_*) between phenotypes. We used default software settings for LDSC analyses and included individuals of matching ancestry from the 1000 Genomes Consortium phase 3v5 (1000 Genomes Project Consortium et al., 2015) as an LD reference panel. Phenotypes with non-calculable or low *h*^2^_SNP_ (Z-score < 3) were excluded from further analysis (**Table S2**), resulting in a final set of 18 AUBs, summarized in **Table 1**.

**Table 1.**
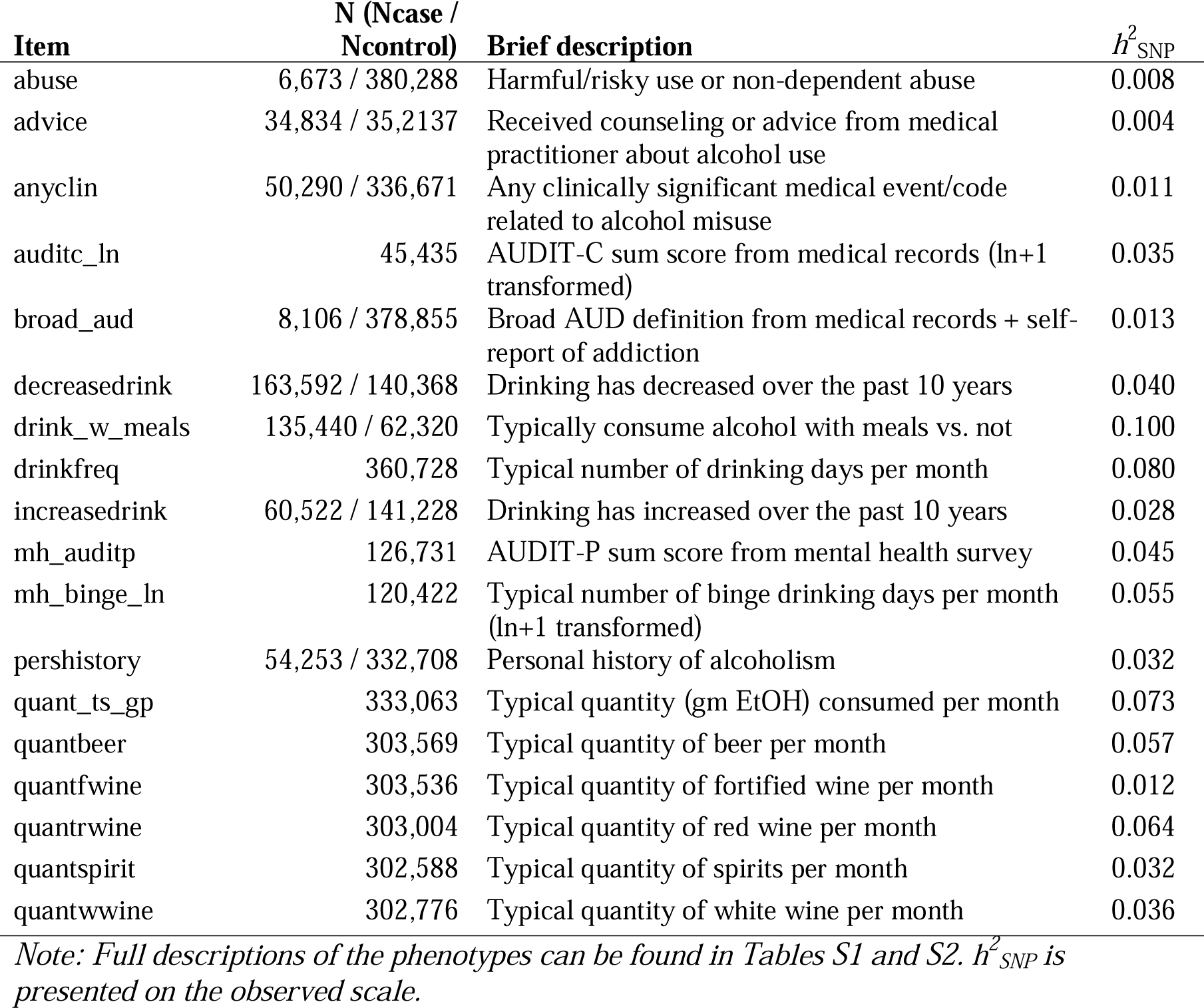
Summary of univariate AUB GWAS phenotypes included in the genetic factor models.

### Structural equation modelling

To empirically model the genetic structure of AUBs, we conducted genomic structural equation modelling (gSEM) on the genetic correlations between the 18 phenotypes using the *GenomicSEM* package v0.0.5 (Grotzinger et al., 2019) in R v4.2.2 (R Core Team, 2017). The sampling covariance matrix was smoothed using the *nearPD* function with a tolerance of 1e-08. We used the *psych* package (Revelle, 2022) to conduct exploratory factor analysis (EFA) with varimax rotation and the *GenomicSEM* package to conduct confirmatory factor analysis (CFA) based on the EFA results. In the CFA models, indicators were retained for each factor with a loading ≥ 0.4, or for the factor with the highest loading if loadings were < 0.4 on all factors. The best CFA model was chosen based on interpretability and fit statistics of 1-to 5-class factor solutions, including the comparative fit index (CFI; values >.9 indicating good fit) and the standardized root mean square residual (SRMR; values <.08 indicating good fit).

### Genetic correlations

gSEM was also used to provide external validation of the latent factors by examining their genetic correlation with a variety of psychiatric, neurocognitive, physiological, and socioeconomic phenotypes from previous well-powered GWASs. The best-fitting gSEM model was modified to include each external phenotype as a predictor of the latent factors.

Consistency of the genetic correlation was tested by constraining the coefficient to equality across latent factors and comparing the change in model χ^2^. Heterogeneity (*Q*_Trait_) tests were used to test whether the observed genetic correlations were consistent across factor indicators.

### Multivariate latent factor GWAS

GWAS of each of the latent factors from the best-fitting model was conducted with gSEM, which uses the genetic covariance between items to identify the effects of single nucleotide polymorphisms (SNPs) on the latent factor(s). Heterogeneity (*Q_SNP_*) tests were performed to investigate whether each SNP showed a consistent effect across all indicators of a latent trait, and gene-based tests of the *Q_SNP_* statistics indicated whether heterogeneous effects clustered within genes. We used multiple downstream *in silico* approaches to interpret the GWAS results of each of the AUB latent factors, including FUMA (Watanabe, Taskesen, van Bochoven, & Posthuma, 2017) to define the associated genomic loci and prioritize implicated genes, MAGMA (de Leeuw, Mooij, Heskes, & Posthuma, 2015) to aggregate the association statistics for individual SNPs into tests of enrichment within protein-coding genes, and a combination of FUMA, MAGMA, and CELLECT (Timshel, Thompson, & Pers, 2020) to test enrichment of association in specific tissues and brain cell types. Full details of these analyses are in the **Supplementary Methods**.

### Trans-ancestry extension

To examine the generalizability of the EUR latent factor model, we extended the latent factor gSEM model to the UKB African (AFR; n=7,827) and South Asian (SAS; n=9,645) ancestry subgroups. We ran the same univariate GWASs and genetic correlations in unrelated individuals from each ancestral group, using the corresponding 1000 Genomes ancestry reference panel. We applied gSEM as above, but using the best-fitting EUR model to designate the number of factors and sets of items loading on each factor. Although sample sizes for these groups are small, trans-ancestry analyses are sorely lacking in the psychiatric genetics field and there is a critical need to determine the extent to which genetic findings (primarily in EUR samples) are generalizable across population groups.

### Out-of-sample predictions

We derived polygenic scores (PGS) for the latent AUB factors using PRS-CS “auto” version (Ge, Chen, Ni, Feng, & Smoller, 2019) in three independent samples: 7,353 individuals from the Collaborative Study on the Genetics of Alcoholism (COGA) study (Begleiter & Reich, 1995), and individuals from two hospital-based biobanks in the US, Vanderbilt University Medical Center (BioVU, n=72,824) and Mass General Brigham (MGBB, n=30,201). PGSs were used individually to predict AUD diagnoses and severity, and in a phenome-wide association (PheWAS) to broadly characterize their association with an array of medical conditions (**Supplementary Methods**).

## Results

### Univariate GWAS

Univariate GWASs were conducted on 18 AUBs (**Table 1**; **Table S1**). Virtually all traits showed significant associations with ethanol metabolizing enzyme genes on chromosome 4 (**Figure S1**). All traits had significant *h^2^_SNP_* estimates, ranging from 0.5% to 14.8%, with mean χ^2^ statistics of 1.037-1.633 (**Table S2**). The LD score regression intercepts (1.004 – 1.095) indicated that inflation is attributable to polygenicity rather than spurious confounding. The genetic correlations varied among traits (*r_g_* =-.826 to .887; **Table S3**).

### Structural equation modelling

EFA was performed on the genetic correlation matrix with 1-to 5-factor solutions (**Table S4**). Comparison indices of the CFA (**Table 2**) showed that a 4-factor solution provided the best fit, with 77% of the variance explained in the EFA. Although the EFA indicated an orthogonal solution, allowing for correlations between the latent factors improved the fit and the 4-factor oblique model produced acceptable fit indices (CFI = 0.982, SRMR = 0.088). In the 5-factor EFA, the variance explained did not increase beyond 77% and only a single item loaded onto the fifth factor, with the CFA failing to converge due to the sparsity of indicators. For these reasons, we selected the 4-factor model (**Figure 1**, **Table S5**). Although some factor loadings dropped below 0.4, removing these indicators substantially decreased fit (CFI = 0.843, SRMR = 0.263), so they were retained in the final model.

**Table 2.** Comparison of model fit indices from confirmatory factor analysis of the genetic correlation structure of alcohol-related items.

| Factors | Chi-square | df | P | AIC | CFI | SRMR | Factor Correlation |
| --- | --- | --- | --- | --- | --- | --- | --- |
| 1 | 244107.20 | 135 | <.0001 | 244179.20 | 0.545 | 0.253 | n/a |
| 2 | 28165.26 | 128 | <.0001 | 28251.26 | 0.948 | 0.133 | Oblique |
| 2 | 36863.94 | 129 | <.0001 | 36947.94 | 0.932 | 0.137 | Orthogonal |
| 3 | 13120.84 | 125 | <.0001 | 13212.84 | 0.976 | 0.137 | Oblique |
| 3 | 44967.81 | 112 | <.0001 | 45049.81 | 0.750 | 0.204 | Orthogonal |
| **4** | **9507.76** | **120** | **<.0001** | **9609.76** | **0.982** | **0.088** | Oblique |
| 4 | 69439.97 | 126 | <.0001 | 69529.97 | 0.871 | 0.212 | Orthogonal |

*Note: \*Results from the 5-factor model are not shown because this model was not identified due to the single item loading on factor 5.*

**Figure 1.**
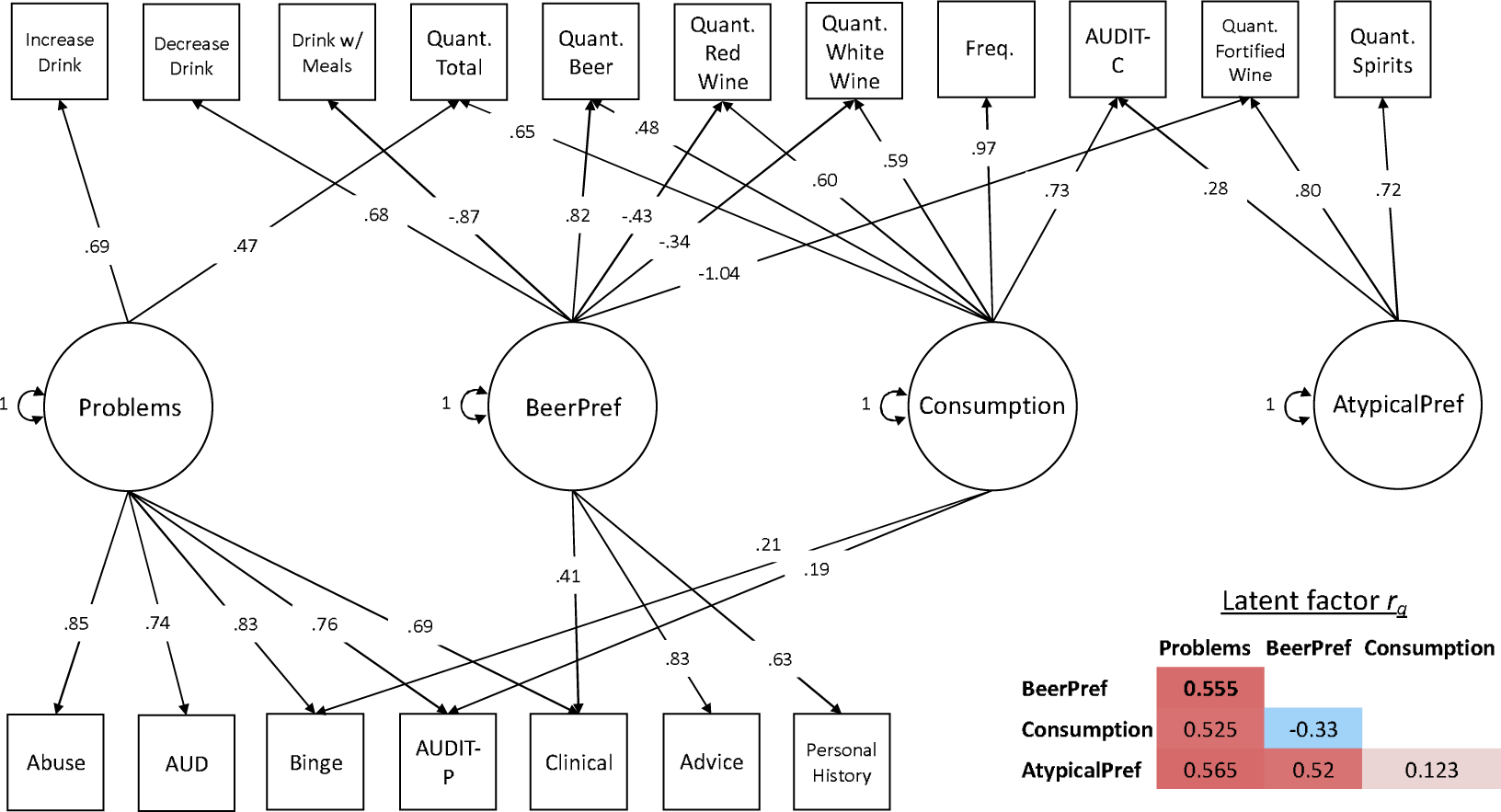
Genetic factor structure of alcohol use behavior phenotypes from the best-fitting model. *Note: Standardized factor loadings are presented. Correlations between latent factors shown in the inset. Full model results shown in Table S5.*

Factor 1 (*Problems*) seemed to capture alcohol-related problems, with strong loadings for items related to misuse: binge drinking and problematic consequences of alcohol use as measured via self-reported questionnaires (AUDIT problems scale scores; AUDIT-P) and clinical diagnoses. Factor 2 (*BeerPref)* reflected a pattern of drinking without meals and drinking beer, specifically, but not other types of alcohol. This factor also indexed some alcohol-related problems, such as receiving advice from a doctor to reduce drinking and experiencing clinically significant consequences, but these went in hand with a pattern of *decreasing* drinking in the past 10 years. Factor 3 (*Consumption*) primarily represented consumption, with strong loadings on AUDIT consumption scale scores (AUDIT-C), overall frequency and quantity of both wine and beer, and weak loadings on binge drinking and AUDIT-P. Finally, Factor 4 (*AtypicalPref*) captured drinking uncommon types of alcohol, such as fortified wine and spirits, with a weaker loading on AUDIT-C.

### Genetic correlations

Genetic correlations between the four factors and an array of external phenotypes are presented in **Figure 2** and **Table S6**. As expected, the strongest correlations were observed for measures of alcohol use or problems, which were consistently positively genetically correlated with *Problems* (*r_g_* > .65) but varied across other factors. For example, the magnitude of the genetic correlation with AD was significantly (*P* < 5.55 × 10^-49^) higher for *BeerPref* (*r_g_* = .56) than for *Consumption* and *AtypicalPref* (*r_g_* = .17-.29), while the magnitude of genetic correlation with AUDIT total scores was significantly (*P* < 8.85 × 10^-20^) higher for *Consumption* (*r_g_* = .85) than all other factors (*r_g_* = .09-.62). *Problems, BeerPref*, and *AtypicalPref* had similar general patterns of correlations with most neuropsychiatric and anthropometric phenotypes, but *BeerPref* showed the strongest correlations with cognitive ability, body size measures, smoking behaviors, and neuropsychiatric outcomes like ADHD and insomnia. For most phenotypes, *Consumption* displayed a pattern of correlations opposite that of the other factors, generally being correlated with *better* physical and mental health.

**Figure 2.**
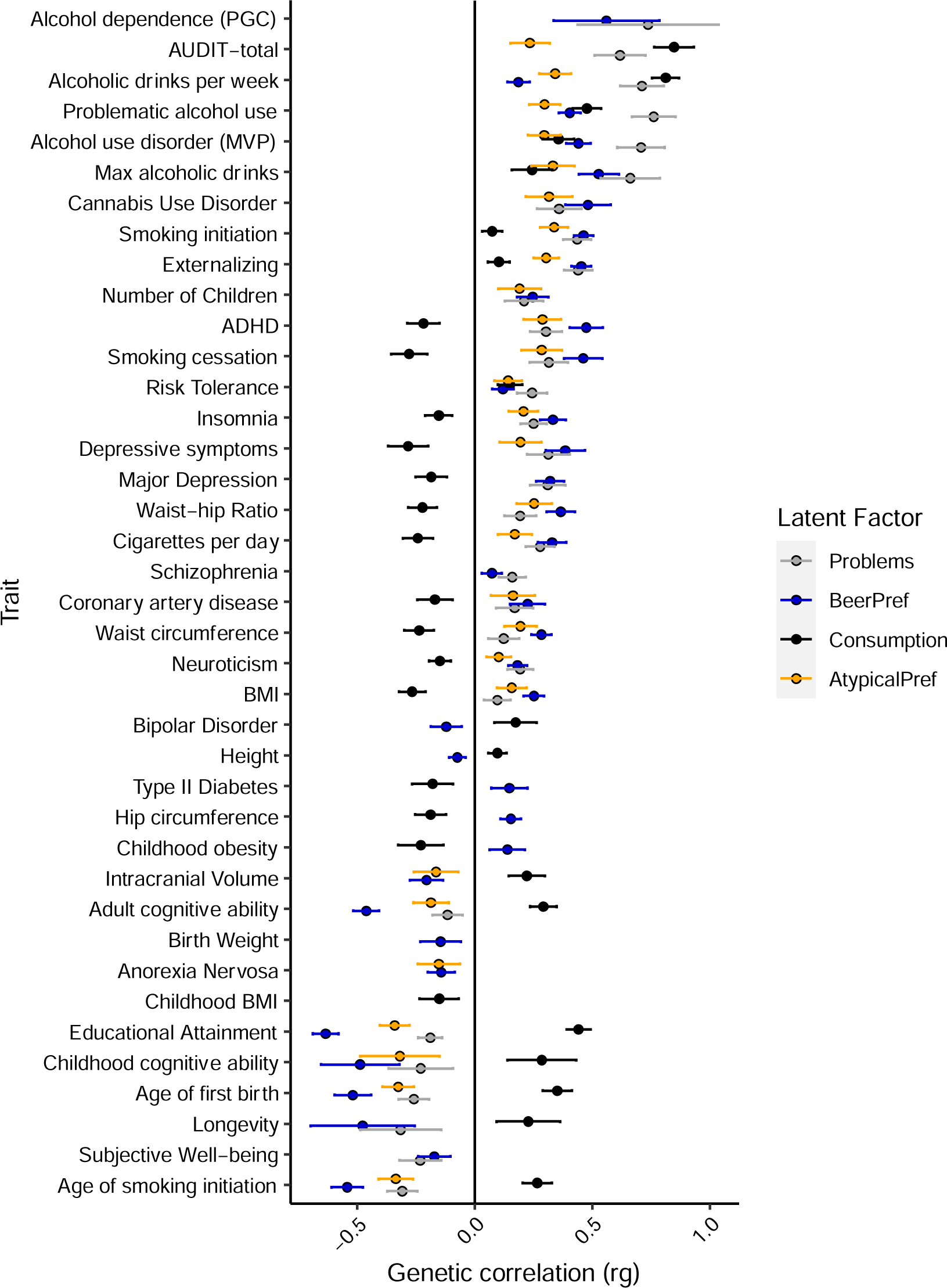
Genetic correlations between the latent factors and external phenotypes. *Note: Results are shown only for significant correlations after correction for 46 correlated phenotypes (p < .001).*

*Q_Trait_*analyses (**Table S6**) indicated that, despite the high factor loadings, there was heterogeneity across the factor indicators in the genetic correlations with approximately one-third of the external phenotypes for *AtypicalPref*, half for *Problems* and *Consumption*, and nearly all external phenotypes for *BeerPref*.

### Multivariate latent factor GWAS

Manhattan plots from the GWASs of these four latent factors are shown in **Figure 3**. Genome-wide independent loci (*r^2^* < .6) are described in **Table S7** and the genes implicated by the GWAS results are in **Table S8** and **S9**. A total of 95 distinct loci were found, which partially overlapped across factors (**Figure S2**). Of those, 50 loci did not overlap regions previously associated with unidimensional AUBs (**Supplementary Methods**).

**Figure 3.**
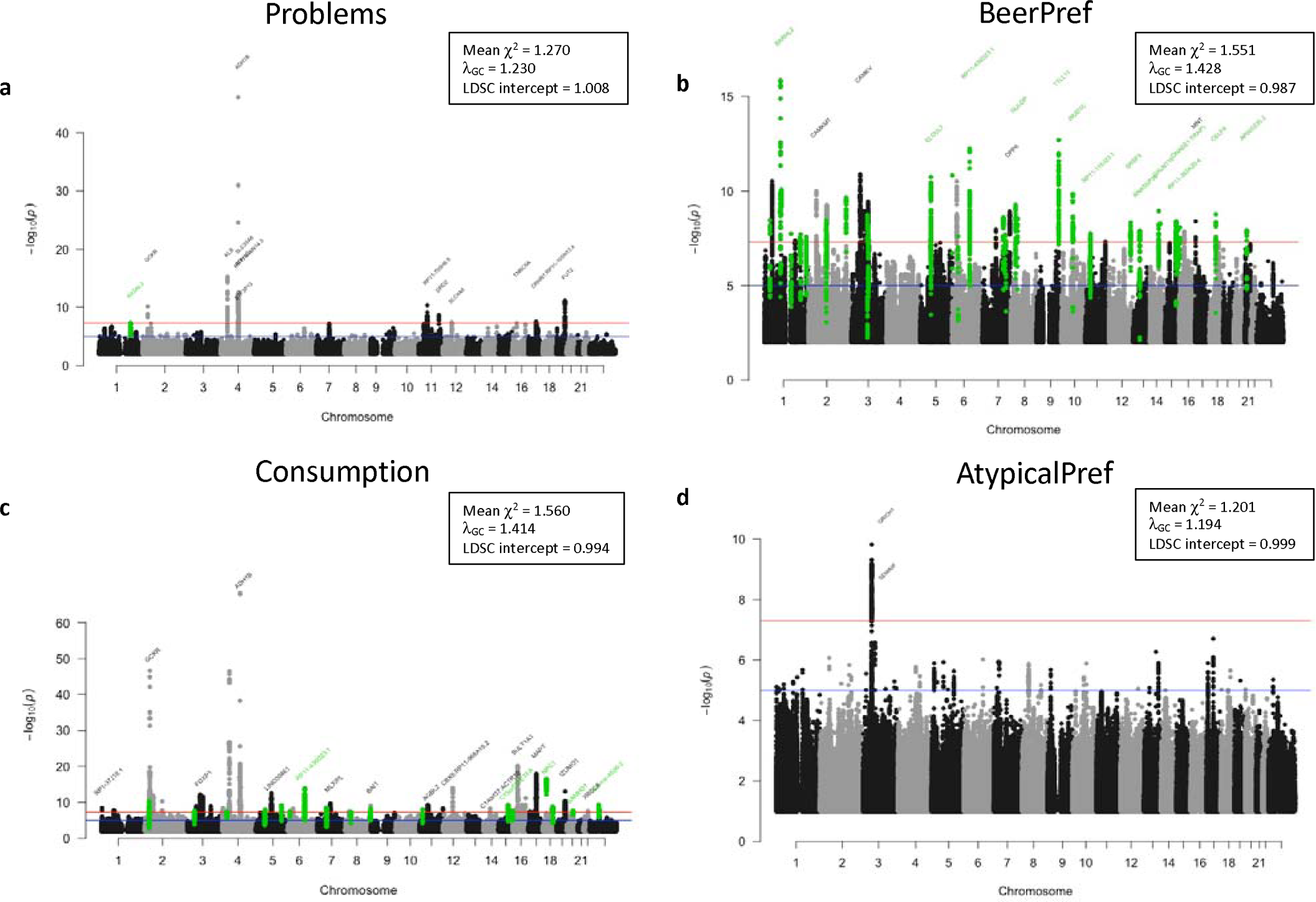
Manhattan plots of the latent factor genome-wide association (GWAS) results. *Note: Results are shown for the four factors represented in Figure 1: a) Problems, b) BeerPref, c) Consumption, and d) AtypicalPref. The dashed line shows the Bonferroni-corrected level of significance (p < 5 × 10^-8^) and the nearest gene is shown for the top locus on each chromosome. Novel loci not identified in previous GWAS of alcohol-related phenotypes are shown in green.*

### Definition and comparison of genomic risk loci

For *Problems*, there were 11 significant loci (**Figure 3a**), including one novel locus on chromosome 1 containing the gene *RASAL2* (chr1:178037791-178449842, *P* = 4.55 × 10^-8^). The strongest signals came from the *ADH1B* (*P* = 7.99 × 10^-47^) and *KLB* (*P* = 5.47 × 10^-16^) gene regions on chromosome 4 (**Table S7**). Heterogeneity (*Q_SNP_*) analyses (**Figure S3**) identified ten genomic regions in which the SNP effects were indicator-specific, most strongly the *ADH1B* locus on chromosome 4 (rs1229984; *P* = 5.19 × 10^-35^), and a variant downstream of *SERTAD2* on chromosome 2 (rs7574806; *P* = 4.49 × 10^-53^). MAGMA gene-based tests indicated that only the gene *CNTNAP5* showed aggregate evidence of heterogeneity in SNP effects between *Problems*-related AUBs (**Figure S4**).

For *BeerPref*, there were 45 significant loci (**Figure 3b**, **Table S7**), of which 32 were novel and 6 overlapped the other factors (**Figure S2**). Surprisingly, SNPs in the *ADH* and *KLB* gene regions on chromosome 4 did not show evidence of association. Instead, novel loci included genes such as *BARHL2* (BarH like homeobox 2) and *TTLL11* (tubulin tyrosine ligase like 11), and strong signal was found in previously identified AUB loci containing calmodulin-related genes (*CAMKV, CAMKMT*). *Q_SNP_* tests showed evidence of heterogeneity at 167 loci, most strongly *ADH1B* (**Figure S3**), as well as other genes with strong links to AUBs in the current and previous studies, such as *CADM2, KLB, STH, FTO, FUT2,* and *GCKR* (**Figure S4**).

For *Consumption*, there were 54 significant loci (**Figure 3c; Table S7**), 18 of which were novel and 12 overlapped with other factors (**Figure S2**). The strongest signals were overlapping the *ADH1B* (*P* = 6.19 × 10^-69^) and *KLB* (*P* = 3.48 × 10^-47^) genes on chromosome 4 and the *GCKR* gene on chromosome 2 (*P* = 2.23 × 10^-47^). Top novel loci included an intergenic region on chromosome 6 (*P* = 1.40 × 10^-14^) and a region including *NPC1* on chromosome 18 (*P* = 3.33 × 10^-17^), an intracellular cholesterol transporter that was also identified in a recent GWAS of multidimensional AUB patterns in the UK Biobank (Thijssen et al., 2023), but not consumption/PAU. *Q_SNP_* tests showed extensive heterogeneity of SNP associations across the genome (**Figure S3**), particularly in genes that were prioritized as top candidates for *BeerPref*, such as *CAMKV, CAMKMT, ELOVL7,* and *TRAIP* (**Figure S4**).

For *AtypicalPref,* there was one significant locus on chromosome 3 (chr3:48564209-50552866, **Figure 3d**, **Table S7**), which overlapped numerous genes and was also associated with *BeerPref*, *Consumption,* and PAU (Zhou et al., 2023). Previous alcohol-related loci such as the *ADH* gene cluster were not significant, although the overall GWAS association signal was low. *Q*_SNP_ analyses showed heterogeneity within *ADH1B* and one locus on chromosome 8 overlapping the genes *TSNARE* and *BAI1*.

### Gene prioritization

Combining four gene-mapping strategies (SNP-based positional, eQTL, and chromatin interaction mapping, and a significant gene-based association; **Tables S8-S9**), there were 19 prioritized genes for *Problems*, 54 for *BeerPref*, 88 for *Consumption*, and 22 for *AtypicalPref* (**Table S10**). These sets of prioritized genes were enriched in numerous previously reported GWAS catalog associations (**Table S11**). For *Problems*, these included neuropsychiatric phenotypes such as PAU, neuroticism and Alzheimer’s disease, as well as anthropometric measures. Prioritized genes for *BeerPref*, *Consumption*, and *AtypicalPref* were enriched for association with phenotypes related to cognition, anthropometrics, brain volume, sleep habits, and social habits (**Table S11**), as well as autoimmune disorders such as ulcerative colitis and Crohn’s disease for *AtypicalPref.* Prioritized genes for *Consumption* also were enriched for expression in the left ventricle of the heart (*P* = 1.32 × 10^-4^) and in middle adulthood (*P* = .004). No other tissue, developmental stage, or gene ontology set was significantly enriched for the other factors.

### Gene-set enrichment

Gene-set analysis was also conducted to investigate enrichment of the SNP-based GWAS association signal in aggregate. For *Problems* there was a significant enrichment in genes highly expressed in the cerebellum and cerebellar hemisphere, and genes with specific expression in the putamen and caudate (**Table S12**, **Figure S5**). For *BeerPref*, there was broad enrichment in genes expressed across all 13 brain regions (*P* < 2.47 × 10^-5^), although not in genes whose expression was specific to any brain region. For *Consumption,* there was enrichment in genes with both high average and region-specific expression in 5 brain regions (cerebellum, cerebellar hemisphere, cortex, frontal cortex, anterior cingulate cortex), in addition to genes with high but not specific expression in the nucleus accumbens, amygdala, caudate, hippocampus, and hypothalamus. For *AtypicalPref*, there was enrichment in genes highly expressed in the cerebellar hemisphere, cerebellum, anterior cingulate cortex, hypothalamus, frontal cortex, caudate, and cortex, and in genes specific to the anterior cingulate and frontal cortex. Overall, the associations from the four latent factor GWASs pointed to genes with functions throughout the brain, and specific regions seem important for all factors except *BeerPref* (cortical regions for *Consumption* and *AtypicalPref*, subcortical for *Problems*).

Gene-set analysis of developmental stage-specific gene expression indicated that, for *BeerPref*, there was enrichment of association signal in genes expressed in early-/mid-prenatal development (*P* = .003) (**Table S12**). There was no significant enrichment in developmental stages for any of the other factors, or in any gene ontology sets.

### Cell type-specific enrichment

Cell type-specific analysis (**Supplementary Methods**) showed a reliable enrichment of SNP association signal for *Problems* in genes expressed in excitatory and inhibitory neurons, particularly upper-layer intratelencephalic neurons in the cortex and medium spiny neurons in the cerebral nuclei (**Table S13**, **Figure S6**). No specific cell types were identified for *BeerPref*. For *Consumption*, excitatory neurons (deep and upper layer interatelencephalic, upper rhombic lip, and amygdala) were reliably enriched for GWAS association signal, as were upper layer intratelencephalic excitatory neurons for *AtypicalPref*.

### Trans-ancestry extension

Factor models within two ancestral groups (AFR, SAS) of the same cohort showed a poor fit to the data (AFR: CFI = .680, SMR = .349; SAS: CFI = .910, SRMR = .261; **Table S14**). The sample sizes for these ancestry groups were small and, consequently, the estimates of the genetic correlations were unreliable.

### Out-of-sample validation

PGS analyses in COGA (*N* = 7,353; mean age [SD] = 37.4 [14.5]; 52.7% female, **Table S15**) demonstrated that each of the latent factors captured genetic risk relevant to clinically diagnosed AUD (**Figure 4a**, left). The four factors captured 1.8%, 1.5%, 0.9%, and 0.3% of the variance in AUD, respectively (3.0% in total). *AtypicalPref* failed to capture significant variance in AUD status after accounting for the other factors (**Figure 4a**, center). PGS of *Problems, BeerPref,* and *Consumption* significantly predicted AUD even after accounting for PGS based on a previous AUD GWAS (**Figure 4a**, right), indicating that these multimodal/multidimensional measures capture relevant genetic risk that is partially independent of unidimensional measures such as AUD. *BeerPref* and *Consumption* were predictive of the full spectrum of AUD severity (mild, moderate, severe), while *Problems* was better able to discriminate severe AUD (**Figure 4b**).

**Figure 4.**
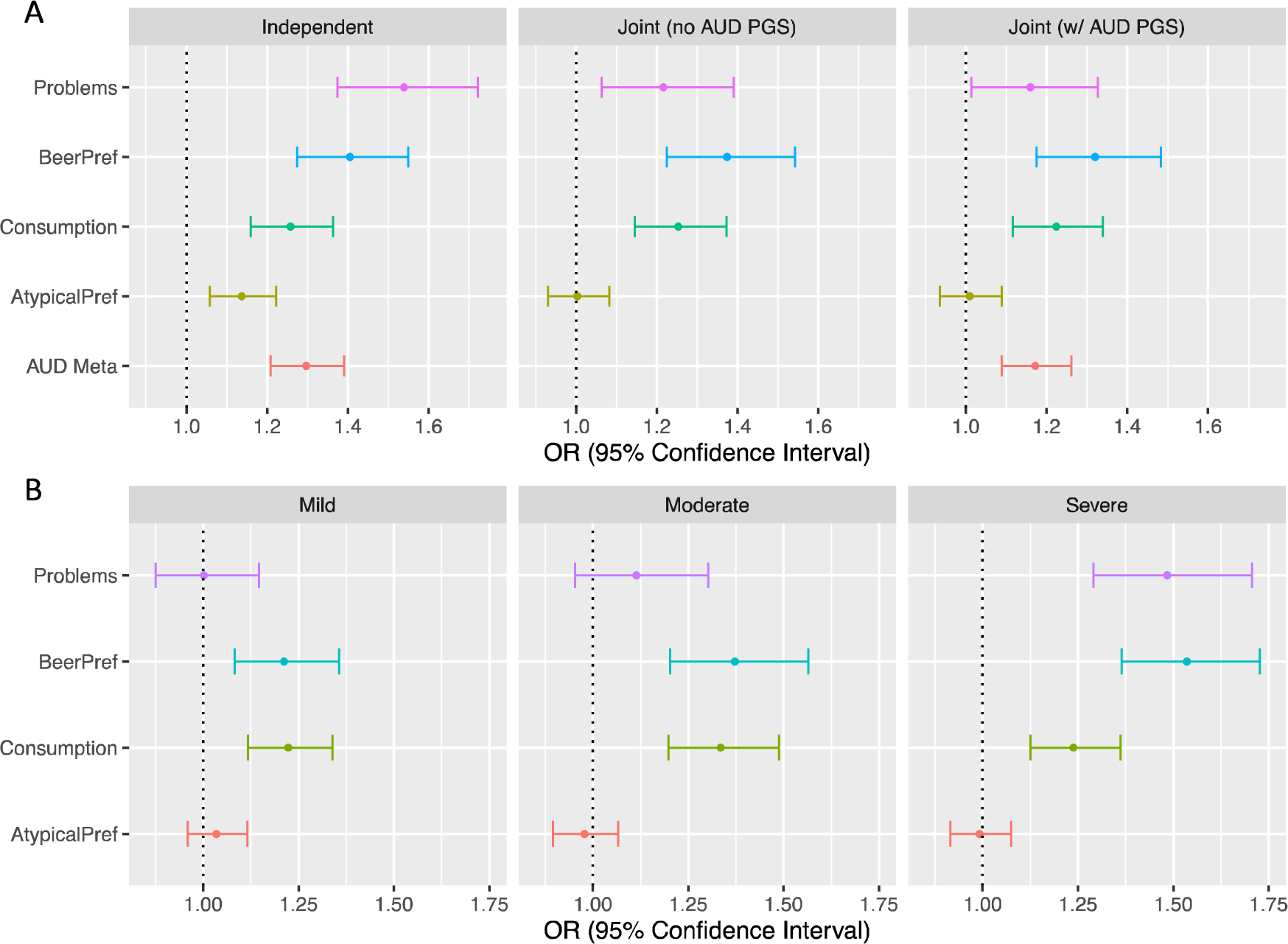
Polygenic score (PGS) prediction of alcohol use disorder (AUD) in an independent sample. *Note: Polygenic scores were derived from the four latent genetic factors represented in Figure 1, as well as a separate GWAS of unidimensional AUD diagnoses (Zhou et al., 2020). (a) Odds ratios (ORs) shown for independent and joint prediction of AUD case/control status by the 5 PGSs. (b) ORs shown for the joint prediction of AUD severity by the 4 latent genetic factor PGSs.*

In the PheWAS analysis in BioVU (**Table S15, Figure S7**), PGS of *Problems* was most predictive of alcohol and other substance use disorders as well as mood and anxiety disorders, while *BeerPref* and *AtypicalPref* most strongly predicted tobacco use and respiratory disorders. *Consumption* was associated with lower risk for cardiovascular disorders, diabetes, and obesity. In the MGBB PheWAS (**Table S16, Figure S8**), all factor PGSs were associated with higher risk of alcohol, substance use, mood, anxiety, and respiratory disorders. *BeerPref* showed particularly strong associations with tobacco use disorder and PTSD, while *Consumption* did not predict lower risk of health conditions in this sample.

## Discussion

In this study we demonstrated that by increasing the resolution of alcohol-related phenotypes, we can improve power for gene discovery and uncover previously unobserved genetic signal relevant to clinical alcohol outcomes. Structural equation modelling and multimodal, multivariate GWAS in a large, population-based sample revealed four latent genetic factors underlying different dimensions of AUBs. These factors in turn were associated with overlapping as well as partially distinct regions of the genome, including 50 that were not identified in prior GWAS of unidimensional AUBs, and which were linked to different profiles of genes, tissues, and cell types.

The structure of the four-factor model is consistent with previous item-level analyses demonstrating a genetic distinction between alcohol problems and alcohol consumption (Kranzler et al., 2019; Mallard et al., 2022; Sanchez-Roige, Palmer, & Clarke, 2020; Sanchez-Roige et al., 2019; Zhou et al., 2020). These two dimensions appeared to be indexed by the latent factors of *Problems* and *Consumption*, respectively, with *Consumption* also replicating a known pattern of genetic correlations between higher consumption and better health outcomes that is likely confounded by socioeconomic status. This may also reflect specific characteristics of this sample such as the known healthy volunteer bias present among participants (Fry et al., 2017), “non-healthy” volunteer bias found in hospital-based biobanks (Feng et al., 2023), as well as possible misreport bias (Xue et al., 2021). Although this confounding was attenuated in previous item-level analyses by separating the unique genetic influences on drinking frequency from drinking quantity (Mallard et al., 2022), the current results suggest that other facets of drinking behaviors, such as the quantity of specific types of alcohol, are influenced by similar confounding processes.

By modeling understudied AUBs, we found evidence of two additional latent factors not seen in previous genetic studies. *BeerPref* appeared to index a number of alcohol-related problems, a pattern of drinking with a specific preference for beer and drinking without meals, alongside (responding to) a doctor’s advice to reduce drinking. The negative genetic correlation between *BeerPref* and *Consumption*, despite both being positively correlated with measures of alcohol use and problems, suggests that these may be capturing distinct risk trajectories with different environmental and sociocultural influences on AUBs. *BeerPref* may also represent genetic risk that is more malleable to intervention, given that it was associated with both receiving advice from a health care practitioner to reduce drinking and a self-reported decrease in recent drinking habits (although the causality between the two cannot be established here). This factor may capture genetic risk for recovering and/or subthreshold AUD – for example, individuals who may have met diagnostic criteria earlier in life but did receive a diagnosis at the time, or who experience related health problems that spur a reduction in drinking (Dao et al., 2021). Genetic risk for such individuals may not be adequately captured in consumption GWASs as current levels of consumption may be lower than during problematic drinking period(s) at other life stages. This issue highlights the need for longitudinal measurement collection to capture dynamic lifelong patterns of AUB, especially given the potential for such changes to bias genetic associations (Xue et al., 2021). Finally, the *AtypicalPref* factor seemed to index consumption of atypical alcoholic beverages such as spirits and fortified wine but was not a very genetically informative factor, likely due to the low prevalence of endorsement of these phenotypes in this cohort.

The extent to which these factors, or individual phenotypes from them, can be used in personalized medicine requires further investigation. There is some support from previous studies that a preference for drinking beer and/or spirits is associated with AUD and overall poorer health outcomes (Niemelä et al., 2022; Smart, 1996). However, these patterns of beverage preferences are extremely difficult to disentangle from confounding socioeconomic and lifestyle factors (Paschall & Lipton, 2005; Sluik, van Lee, Geelen, & Feskens, 2014).

Still, the differential pattern of genetic associations and correlations displayed by the latent factors suggests that beverage preference and habitual patterns of use may be useful indices to identify individuals with different types of risk, even if the causal processes underlying those differences are not yet fully understood. We note that the relevance of different AUBs may be specific to the particular sociocultural context within the UKB European ancestry sample, and other behaviors besides beverage preference may be more relevant to genetic studies in different contexts.

These results suggest that partially distinct genetic factors underlie different AUB dimensions. Although there was moderate genetic overlap across factors, the *Problems* and *Consumption* GWASs largely identified genomic loci with known associations, whereas the *BeerPref* factor identified primarily novel loci. Including more diverse measures of alcohol use/misuse did also boost gene discovery, as novel loci were also identified for both *Problems* and *Consumption*. Notably, one novel locus for *Problems* overlaps the gene *RASAL2*, which is also the only gene in this locus mapped by all 4 methods*. RASAL2*’s protein contains a domain of GTPase-activating proteins which activate Ras, and this gene is involved in response to glucose and has previously been linked to body size and body fat measures, asthma, and the AUDIT item of experiencing memory loss due to drinking (Watanabe et al., 2019). These results highlight *RASAL2* as a candidate for alcohol problems that is not shared with other dimensions of AUBs.

Although further experimental investigations are needed, results indicate that some tissues and cell types (cerebellum, cortex, excitatory neurons) are broadly involved across multiple dimensions of AUBs, while others are more specific in their associations (e.g., caudate, putamen, and inhibitory neurons in alcohol problems). The distinct pattern of enrichment seen for *BeerPref*, with broad associations in brain tissues but no specific regions or cell types, suggests that this latent factor differs qualitatively from other AUBs and merits further investigation. *Q_SNP_* and *Q_T_*_rait_ analyses indicate substantial heterogeneity across specific measures, especially within *BeerPref* and *Consumption* factors, which may also be relevant to individual risk and resilience.

In summary, we have identified multiple distinct genetic factors underlying AUBs, which are both correlated with and predictive of clinically relevant alcohol outcomes. Our analyses provide a large set of prioritized candidate genes for functional follow-up, as well as insight into the genetic architecture of different dimensions of AUBs. Although future research is needed to derive further insight into the biological interpretation of these dimensions, particularly across diverse ancestral groups, our results highlight the promise of deep phenotyping and multimodal, multidimensional assessment to aid our understanding of the etiology of alcohol use behaviors.

### Disclosures

J.W.S. is a member of the Leon Levy Foundation Neuroscience Advisory Board, the Scientific Advisory Board of Sensorium Therapeutics (with equity), has received grant support from Biogen Inc, and is principal investigator of a collaborative study of the genetics of depression and bipolar disorder sponsored by 23andMe for which 23andMe provides analysis time as in-kind support but no payments. The authors report no other financial relationships with commercial interests.

## Acknowledgments

*Primary data analysis:* This research was funded by a grant to J.E.S. (VENI 201G-064) from The Netherlands Organization for Scientific Research (NWO). The research has been conducted using the UK Biobank Resource (application no. 16406). This work uses data provided by patients and collected by the National Health Services England as part of their care and support, as well as data assets made available by National Safe Haven as part of the Data and Connectivity National Core Study, led by Health Data Research UK in partnership with the Office for National Statistics and funded by UK Research and Innovation (MC_PC_20058). We would like to thank the many UK Biobank participants and staff.

*COGA, MGBB, BioVU validation samples:* The Collaborative Study on the Genetics of Alcoholism (COGA), Principal Investigators B. Porjesz, V. Hesselbrock, T. Foroud; Scientific Director, A. Agrawal; Translational Director, D. Dick, includes ten different centers: University of Connecticut (V. Hesselbrock); Indiana University (H.J. Edenberg, T. Foroud, Y. Liu, M.H. Plawecki); University of Iowa Carver College of Medicine (S. Kuperman, J. Kramer); SUNY Downstate Health Sciences University (B. Porjesz, J. Meyers, C. Kamarajan, A. Pandey); Washington University in St. Louis (L. Bierut, J. Rice, K. Bucholz, A. Agrawal); University of California at San Diego (M. Schuckit); Rutgers University (J. Tischfield, D. Dick, R. Hart, J. Salvatore); The Children’s Hospital of Philadelphia, University of Pennsylvania (L. Almasy); Icahn School of Medicine at Mount Sinai (A. Goate, P. Slesinger); and Howard University (D. Scott). Other COGA collaborators include: L. Bauer (University of Connecticut); J. Nurnberger Jr., L. Wetherill, X., Xuei, D. Lai, S. O’Connor, (Indiana University); G. Chan (University of Iowa; University of Connecticut); D.B. Chorlian, J. Zhang, P. Barr, S. Kinreich, G. Pandey (SUNY Downstate); N. Mullins (Icahn School of Medicine at Mount Sinai); A. Anokhin, S. Hartz, E. Johnson, V. McCutcheon, S. Saccone (Washington University); J. Moore, F. Aliev, Z. Pang, S. Kuo (Rutgers University); A. Merikangas (The Children’s Hospital of Philadelphia and University of Pennsylvania); H. Chin and A. Parsian are the NIAAA Staff Collaborators. We continue to be inspired by our memories of Henri Begleiter and Theodore Reich, founding PI and Co-PI of COGA, and also owe a debt of gratitude to other past organizers of COGA, including Ting-Kai Li, P. Michael Conneally, Raymond Crowe, and Wendy Reich, for their critical contributions. This national collaborative study is supported by NIH Grant U10AA008401 from the National Institute on Alcohol Abuse and Alcoholism (NIAAA) and the National Institute on Drug Abuse (NIDA). We thank Mass General Brigham Biobank (MGBB) and Vanderbilt University Medical Center Biobank (BioVU) for providing genomic and health information data. This study would not be possible without the contributions of patients and Biobank participants. We would also like to thank the research coordinators and the Biobank study for their tremendous effort in participant recruitment and sample collection. Lastly, we would like to acknowledge the MGBB Research Patient Data Registry team for their work maintaining the enterprise research patient data warehouse.

*Secondary analyses:* included data from the Million Veteran Program, Office of Research and Development, Veterans Health Administration, and was supported by the Veterans Administration (VA) Million Veteran Program (MVP). The authors thank the staff, researchers, and volunteers, who have contributed to MVP, and especially participants who previously served their country in the military and now generously agreed to enroll in the study. (See https://www.research.va.gov/mvp/ for more details). The citation for MVP is Gaziano, J.M. et al. Million Veteran Program: A mega-biobank to study genetic influences on health and disease. J Clin Epidemiol 70, 214-23 (2016).

Analyses were carried out on the Genetic Cluster Computer hosted by the Dutch National computing and Networking Services SURFsara. The content is solely the responsibility of the authors and does not necessarily represent the views of the funding agencies. The funding agencies had no role in the study design, data analysis, manuscript preparation, or decision to submit for publication.

## Data Availability Statement

Data was obtained from previously collected biobanks whose raw data is available to qualified researchers upon request.

UK Biobank: https://www.ukbiobank.ac.uk/

COGA: https://cogastudy.org/

MGBB: https://www.massgeneralbrigham.org/en/research-and-innovation/participate-in-research/biobank/for-researchers

BioVU: https://victr.vumc.org/biovu-description/

Genome-wide summary statistics will be made publicly available via https://ctg.cncr.nl/software/summary_statistics/ upon publication.

