## Supplementary Methods; Supplementary Figures S1-S7 for "Genetic Heterogeneity Across Dimensions of Alcohol Use Behaviors"

#### Multivariate latent factor GWAS

##### Definition of genomic risk loci

Functional annotation of the AUB latent factor GWAS was performed using the online platform FUMA v1.6.0 (Watanabe, Taskesen, van Bochoven, & Posthuma, 2017), which uses LD patterns to define independent (*r*^2^ < .1) genomic risk loci that reach a genome-wide significant level of association (*p* < 5 × 10^-8^). Candidate SNPs within these loci are annotated with information about known functional consequences from bioinformatics databases and are mapped to genes using the SNP2GENE analysis based on a) genomic position, b) effects on gene expression (expression quantitative loci [eQTLs]), and c) 3-dimensional chromatin interactions. Results of these annotations are available online at <https://fuma.ctglab.nl> (Job IDs: 281242, 281097, 273720, and 273719).

##### Comparison with previous AUB GWAS loci

Associated loci, as defined above, were compared against previous large-scale GWASs of multiple unidimensional AUBs, primarily PAU and consumption (Kranzler et al., 2019; Liu et al., 2019; Mallard et al., 2022; Sanchez-Roige et al., 2019; Walters et al., 2018; Zhou et al., 2023; Zhou et al., 2020). We extracted previously associated loci from the genomic boundaries defined in these prior publications. If only lead SNPs rather than locus boundaries were provided, we defined the locus as a 250kb region centering on the lead SNP. Overlapping or nearby (within 250kb) regions were merged across studies to form a set of 152 “known” alcohol-associated loci whose genomic positions were compared with those of the latent factor-associated loci.

##### Gene-based association testing

MAGMA (de Leeuw, Mooij, Heskes, & Posthuma, 2015) was used to aggregate the GWAS association statistics for individual SNPs into tests of enrichment within protein-coding genes using default program settings. The Bonferroni corrected significance threshold was *p* <.05/19,471 genes = 2.56 × 10^-6.^

##### Gene prioritization

Genes with a significant gene-based association statistic in MAGMA, in addition to being implicated by all 3 SNP-based mapping strategies in FUMA (positional, eQTL, and chromatin interaction) were prioritized as the strongest candidate genes from each latent factor GWAS. We used the GENE2FUNC analysis in FUMA to test for enrichment of these prioritized candidate genes in gene ontology pathways from the Molecular Signatures Database (MSigDb) (Liberzon et al., 2011), tissue-specific expression from the Gene-Tissue Expression Consortium database (GTEx Consortium, 2015), developmental stage-specific expression from BrainSpan, and in previously reported associations with phenotypes in the NHGRI GWAS catalog (MacArthur et al., 2017). GENE2FUNC uses a hypergeometric test to assess whether the set of candidate genes is overrepresented among the defined gene-set genes in comparison to a same-sized set drawn at random from the background pool of protein-coding genes.

##### Gene-set enrichment

MAGMA gene-set analyses were also used to test whether the GWAS association signal was enriched in genes with high/specific expression across tissues and developmental stages, or in genes belonging to known biological pathways. This is similar to the analysis performed on the prioritized genes, described above, but using the full genome-wide SNP-based association values from all genes rather than selecting only the prioritized genes. We computed average gene expression and expression specificity (top 10% of genes expressed specifically in a given cell type) from bulk tissue RNA sequencing for each of 54 tissues in GTEx (GTEx Consortium, 2015). Similar RNA sequencing data from the BrainSpan database was used to identify gene expression patterns at 11 different stages of human development. Gene ontology gene-sets were also obtained from MSigDb (Liberzon et al., 2011). In MAGMA, Bonferroni correction was applied for the number of datasets tested (tissues: p < .05/54=9.26 × 10^-4.^; development: p < .05/11 stages = .0045; gene ontology: p < .05/ 5917 sets=8.45 × 10^-6^).

##### Cell type-specific enrichment

To evaluate whether and which brain cell types are implicated by the aggregate genome-wide association signal from GWAS, we implemented cell type enrichment analyses. We used single cell RNA sequencing (scRNAseq) data from 10 adult human brain regions (Siletti et al., 2022), with multiple dissection datasets within the 10 brain regions serving as a built-in replication test. A count matrix of scRNAseq data was downloaded from the CellxGene portal (<https://cellxgene.cziscience.com/>) on May 12, 2023. Because there are many methods with different underlying statistical methodologies for linking GWAS to cell type-specific gene enrichment, we implemented three methods that span multiple ways of summarizing the scRNAseq data (specifically mean and specificity) and different statistical methodology (MAGMA and LDSC). We looked for cross-method convergence with the hypothesis that if a cell type is predicted to be significant by multiple methods and multiple datasets, it is more likely to be associated with a trait.

Specifically, we used MAGMA to test for cell type enrichment using mean gene expression per cell type as implemented in the FUMA platform by Watanabe, Umićević Mirkov, de Leeuw, van den Heuvel, and Posthuma (2019), hereafter referred to as FUMA Cell type. Another method to link cell types and GWAS is MAGMA_Celltyping as implemented by Skene and Grant (2016). With this method, instead of mean gene expression, we computed the top 10% of genes specific to each cell type calculated using Expression Weighted Celltype Enrichment EWCE). Finally we implemented CELLECT where each cell-type’s Expression Specificity profile was calculated with CELLEX (Timshel, Thompson, & Pers, 2020). Within CELLECT, we used both approaches for calculating the specificity profiles: MAGMA (hereafter referred to as cellect_magma) and LDSC (hereafter referred to as cellect_ldsc). In total, we carried out the single-cell enrichment analysis across 30 cell types using 4 methods. Results were considered credible if there was a significant association in >3 datasets and at least 2 methods, after Bonferroni correction for the number of cell types tested within each method. Although using multiple approaches gives less clarity than a single test statistic, there is currently much debate in the field around the interpretation of different methods and there is no best practice established (Olislagers, Rademaker, Adan, Lin, & Luykx, 2022), so we chose to draw general conclusions from the results that seem most replicable and robust.

#### Out-of-sample predictions

We carried out polygenic score (PGS) prediction in three independent samples: 7,353 individuals from the Collaborative Study on the Genetics of Alcoholism (COGA) study (Begleiter & Reich, 1995), and individuals from two hospital-based biobanks in the US, Vanderbilt University Medical Center (BioVU, n=72,824) and Mass General Brigham (MGBB, n=30,201). Specific details about each of these samples are below.

##### COGA

COGA ascertained high-risk families through adult probands in treatment for alcohol dependence, beginning in 1989. Recruitment was extended to include additional relatives and community-ascertained comparison families (Total *N* = 16,848). Data collection included a psychiatric interview (the Semi-Structured Assessment for the Genetics of Alcoholism, or SSAGA; (Bucholz et al., 1994)). Genotyping was conducted across different phases of data collection. European ancestry (EA) samples were genotyped across multiple arrays, including: (1) the Illumina HumanHap1M array; (2) the Illumina OmniExpress; and (3) the Affymetrix Smokescreen array. Principal components were computed from GWAS data using Eigenstrat (Price et al., 2006) and 1000 Genomes, Phase III reference panel (1000 Genomes Project Consortium et al., 2015). For all analyses within COGA, we included age (of last interview), biological sex, genotyping array, collection site, and the first 10 ancestral PCs as covariates. We corrected standard errors within families using a cluster-robust estimator (Cameron, Gelbach, & Miller, 2011).

##### BioVU

We used de-identified clinical data from individuals in BioVU (Roden et al., 2008). Genotype data were generated using the Illumina Multi-Ethnic Genotype Array (MEGAEX) for 72,824 individuals. Details on the quality control process have been described elsewhere (Dennis et al., 2021). Genotypes were filtered for SNP (<0.95) and individual (<0.98) call rates, sex discrepancies, and excessive heterozygosity (|Fhet|>0.2).82 The sample was then filtered for cryptic relatedness by removing one individual of each pair for which pihat>0.2. PCA using FlashPCA2 (Abraham, Qiu, & Inouye, 2017) combined with CEU, YRI and CHB reference sets from the 1000 Genomes Project Phase 3 (1000 Genomes Project Consortium et al., 2015). was implemented to determine European ancestry. We confirmed the absence of genotyping batch effects. We imputed genotypes using the Michigan Imputation Server with the reference panel from the Haplotype Reference Consortium (McCarthy et al., 2016). SNPs were filtered for imputation quality (R2 >0.3 or INFO >0.95) and converted to hard calls. We restricted the analyses to autosomal SNPs with minor allele frequency (MAF)<0.01. We removed SNPs that differed by >10% from the 1000 Genomes Project phase 3 CEU set and those with a Hardy Weinberg Equilibrium (HWE) p<1.00E-10. The resulting data set contained hard-called SNP information for 9,386,383 SNPs in 72,824 individuals of European Ancestry.

##### MGBB

Samples from MGBB (Boutin et al., 2022) were genotyped in two batches. The first subset of MGB Biobank samples (N_Initial, MEGA_ = 36,424; hereafter, MEGA samples) were genotyped on Multi-Ethnic Global Array (MEGA) kits from Illumina (Illumina Inc., San Diego, USA) and released in eight batches. We performed batch-specific genotype data QC to remove single nucleotide polymorphisms (SNPs) with genotype missing rate >0.05, samples with genotype missing rate >0.02, and SNPs with differential missing rate >0.01 between any two batches, after which different batches were merged for subsequent QC steps.

As MGB Biobank included individuals from diverse populations, we inferred the genetic ancestry of biobank participants using 1000 Genomes samples (1KG) as the population reference panel (1000 Genomes Project Consortium et al., 2015). Specifically, we computed principal components (PCs) for biobank samples and 1KG samples combined and trained a random forest classifier to assign a “super population” label for biobank samples with a prediction probability ≥0.9 using the first 6 PCs of the 1KG samples as the training data. This resulted in 25,677 individuals whose ancestry was classified as European (EUR), 1,607 as African (AFR), 1,840 as Admixed American (AMR), 504 as East Asian (EAS), and 297 as South Asian (SAS) ancestry.

The second subset of MGB Biobank samples (N_Initial, GSA_ = 47,321; hereafter, GSA samples) were genotyped on Global Screening Array (GSA) kits from Illumina (Illumina Inc., San Diego, USA) and released in three batches. We performed batch-specific genotype data QC and computed PCs using the same criteria and procedures as was done in the MEGA samples. This resulted in 33,614 individuals whose ancestry was classified as EUR, 2,036 as AFR, 2,599 as AMR, 908 as EAS, and 480 as SAS ancestry.

The following procedures were applied to both MEGA and GSA samples. Within each ancestry, we excluded samples with mismatched reported and genetic sex, outliers of the absolute value of heterozygosity (>5 standard deviations from the mean), and one sample from each pair of related individuals (identity-by-descent (IBD) >0.2); SNPs that showed significant batch associations at P < 1 × 10−4, had a missing rate > 0.02 or Hardy–Weinberg equilibrium (HWE) test P < 1 × 10−10 were also discarded. Next, we used the Michigan Imputation Server (Minimac4) to impute genotype dosages for biobank samples, with the Haplotype Reference Consortium (HRC) as the reference panel for EUR ancestry. Lastly, we removed markers with imputation quality INFO score <0.8, minor allele frequency (MAF) <0.01, a significant deviation from HWE with P < 1 × 10−10, and missing rate >0.02. The dataset uses genome build 37 (hg19). Further information about genotyping, QC, imputation, and population assignment procedures for the MGB Biobank is available on the GitHub repositories for the respective samples (MEGA samples: <https://github.com/Annefeng/PBK-QC-pipeline>, GSA samples: <https://github.com/getian107/MGBB-QC>).

In this study sample of 30,201 participants, 14,987 participants had DNA genotyped using both MEGA and GSA kits, resulting in duplicate genotype data. With MEGA providing a denser array with approximately 1.7 million variants as opposed to the GSA’s coverage of around 660,000 variants, we prioritized the MEGA samples for their broader variant coverage (Martin et al., 2021).

##### PGS prediction

In all three samples, we derived polygenic scores (PGS) for the latent AUB factors using PRS-CS “auto” version (Ge, Chen, Ni, Feng, & Smoller, 2019). In COGA, we tested the association between AUB latent factor PGS and DSM diagnoses of overall AUD and AUD severity (mild, moderate, severe). PGS for each of the latent factors were used a) individually, b) jointly, and c) jointly alongside a PGS of AUD (Zhou et al., 2020) to identify the unique contributions of each latent factor and determine whether the latent factors capture genetic risk for clinically significant alcohol problems over and above unidimensional GWAS measures. In BioVU and MGBB, we extended the PGS analyses to a full phenome-wide association (PheWAS) to test the association between AUB latent factors and ICD diagnoses for AUD and an array of medical conditions and diagnoses. We performed PheWAS by fitting logistic regression models to predict case/control status across numerous diagnostic codes (“phecodes”; https://phewascatalog.org/phecodes) in electronic health records which have previously been categorized into nosologically similar groups based on their shared etiology or clinical presentation. Analyses were conducted using the PheWAS v0.12 R package (Carroll, Bastarache, & Denny, 2014) adjusting for sex, median age and the first ten PCs of genetic ancestry. Bonferroni correction was applied for each site-specific analysis to account for multiple testing (BioVU: p < .05/1343 phecodes=3.74 × 10^-5^; MGBB: p < .05/1762 phecodes=2.84 × 10^-5^).

### SUPPLEMENTARY FIGURES

##### Supplementary Figure S1. Manhattan plots of GWAS association results for 18 alcohol-related behaviors used in genomic structural equation modeling.

abuse	advice

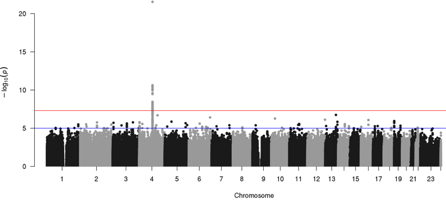

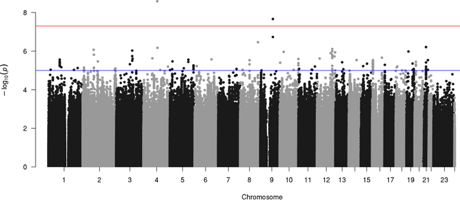

anyclin	auditc_ln

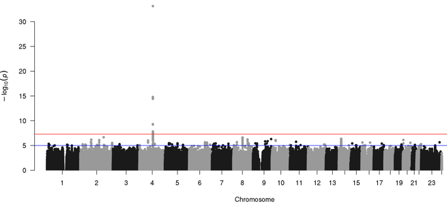

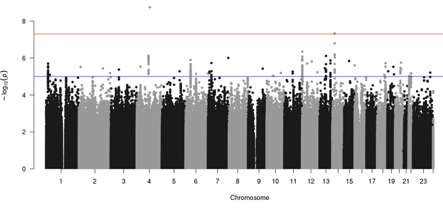

broad_aud	decreasedrink

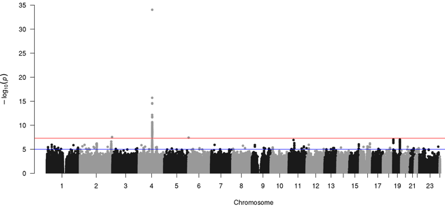

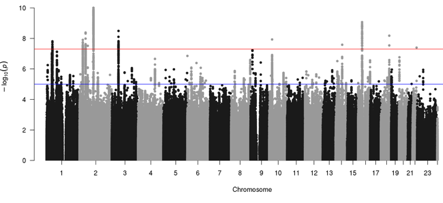

drink_w_meals	drinkfreq

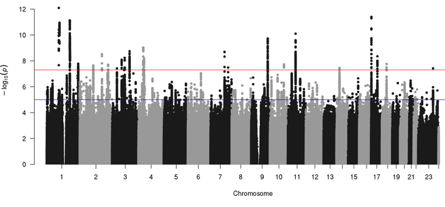

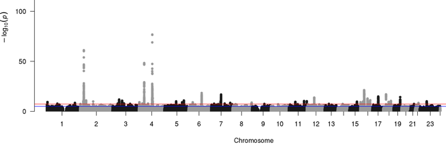

increasedrink	mh_auditp

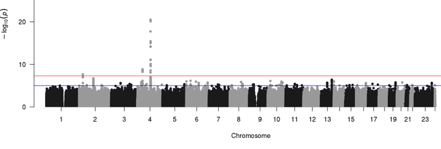

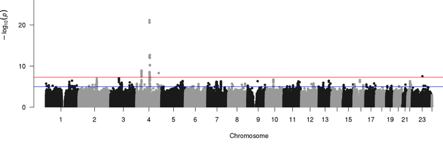

mh_binge_ln	pershistory

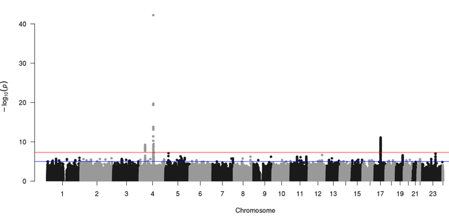

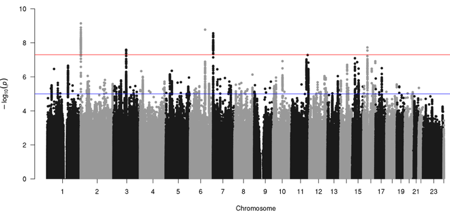

quantbeer	quantfwine

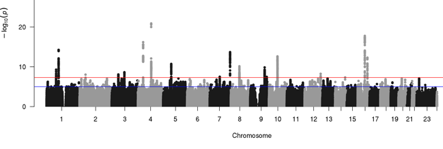

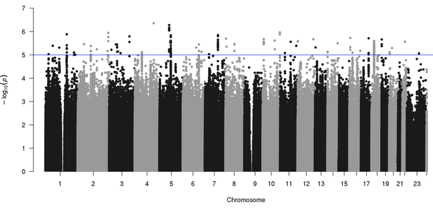

quantrwine	quant_ts_gp

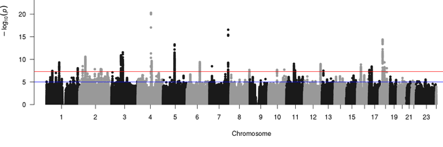

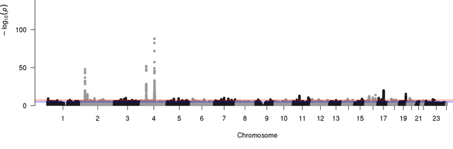

quantwwine	quantspirit

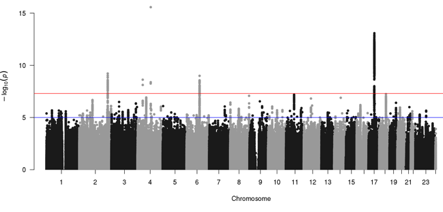

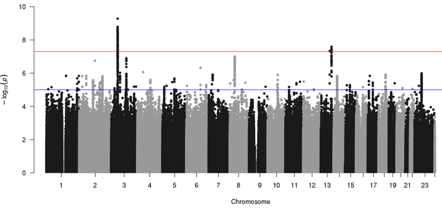

##### Supplementary Figure S2. Unique and overlapping genetic loci across latent factors and previous known loci linked to alcohol use behaviors.

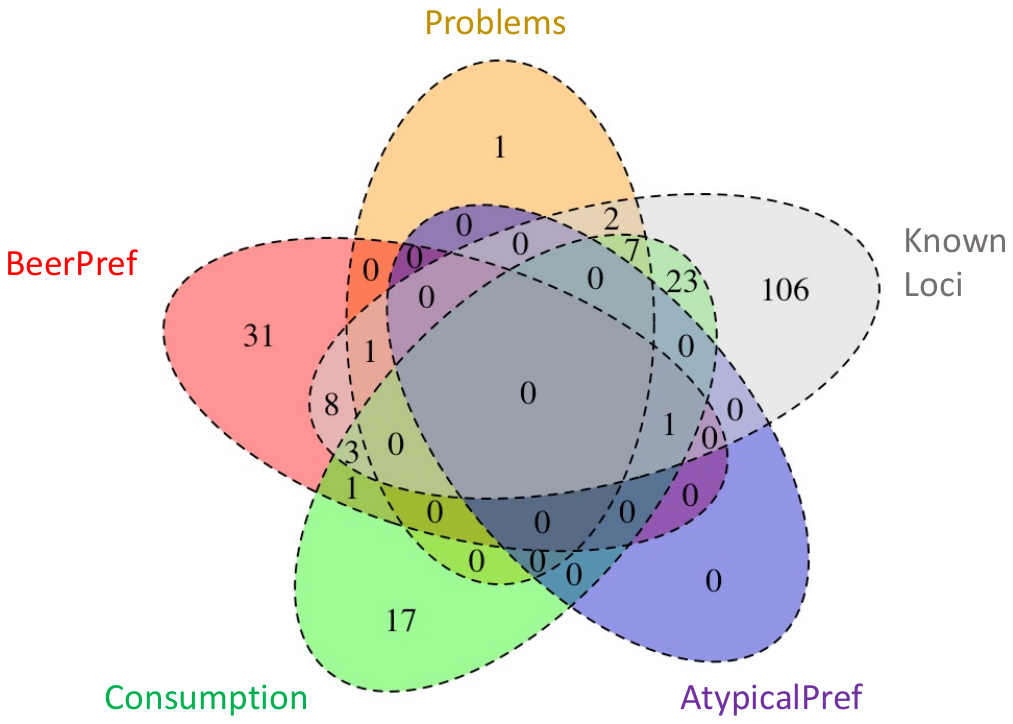

##### Supplementary Figure S3. Heterogeneity (Q_SNP_) of genetic variant associations across indicators of the latent factors.

Problems

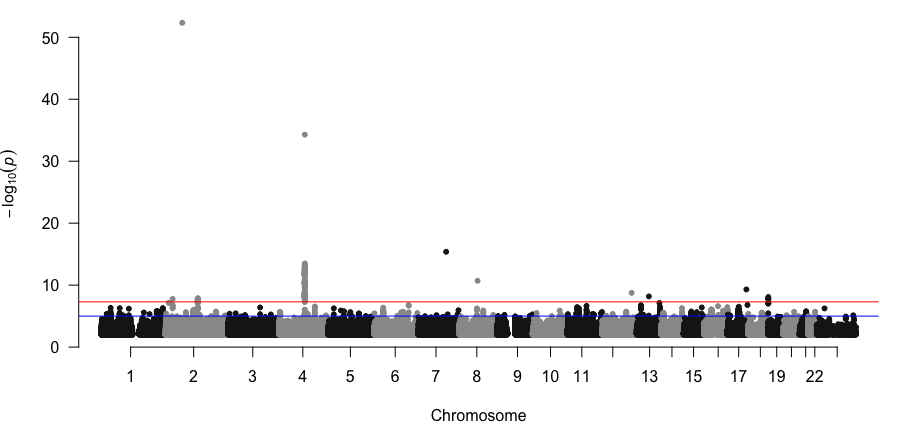

BeerPref

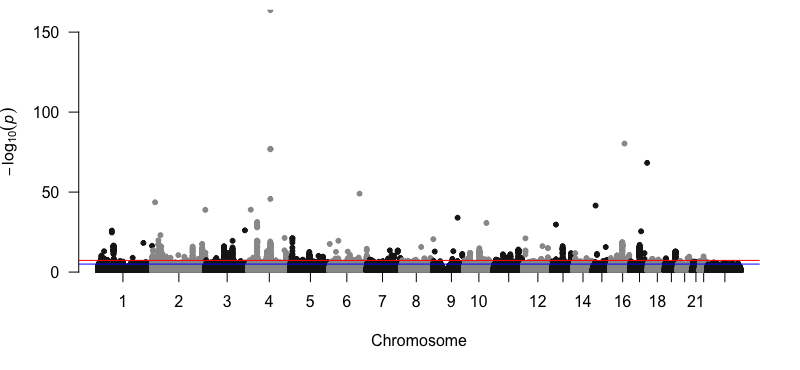

Consumption

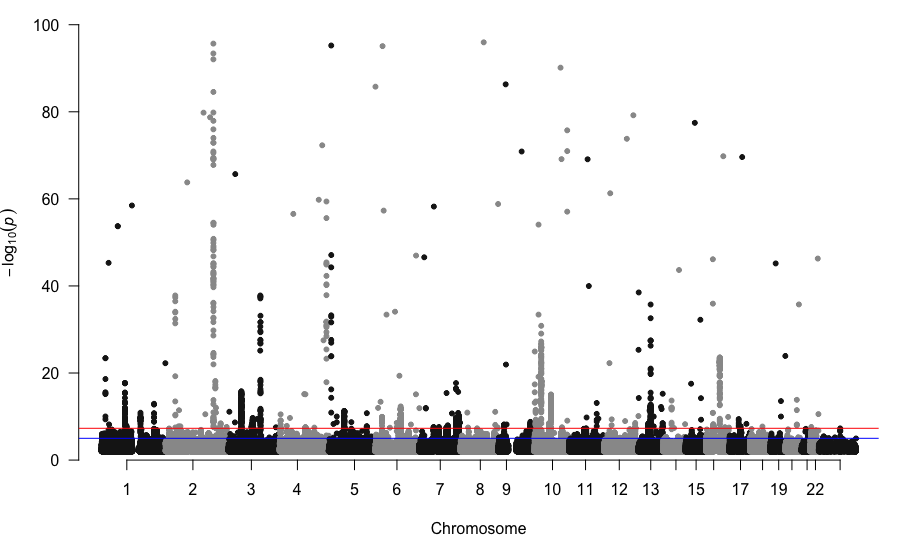

AtypicalPref
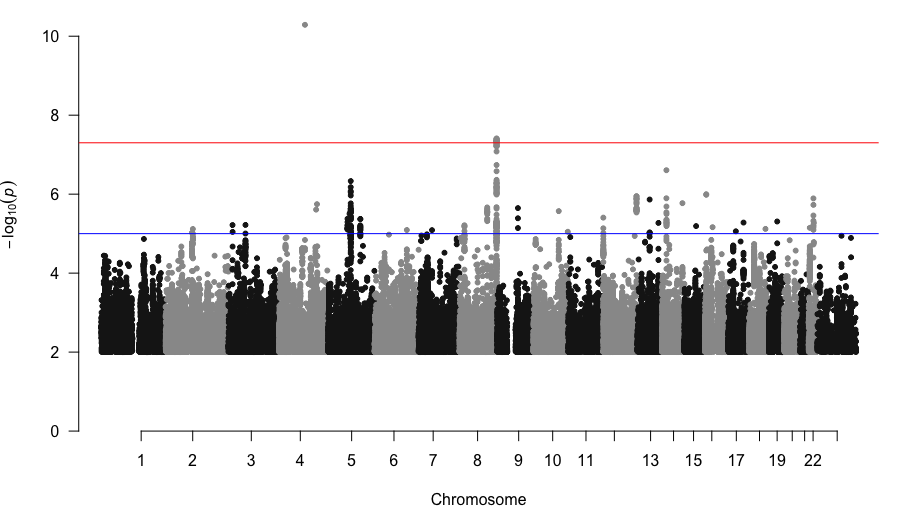

##### Supplementary Figure S4. MAGMA gene-based aggregation of SNP heterogeneity (Q_SNP_) test results for the latent factors.

Problems

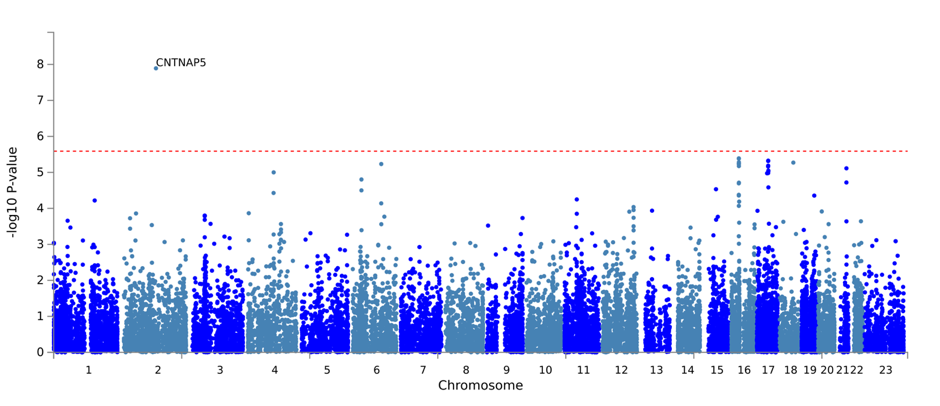

BeerPref

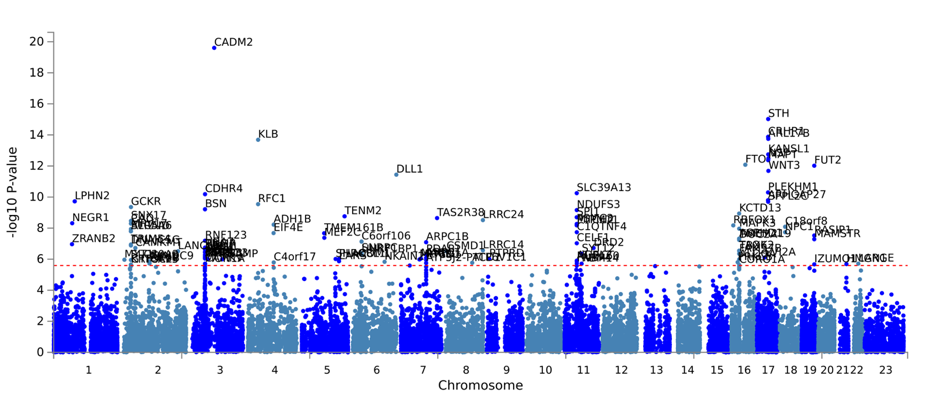

Consumption

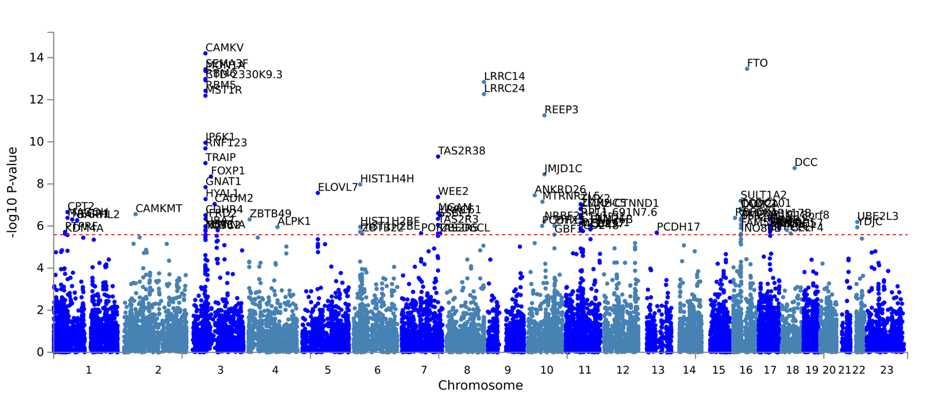

AtypicalPref

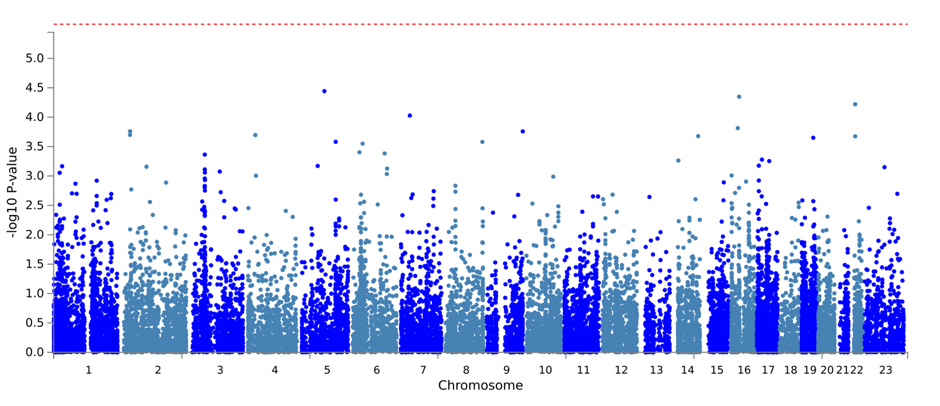

##### Supplementary Figure S5. Enrichment of GWAS association signal for the latent factors in genes with high average expression (MeanLinear) and specific expression (SpecTop10) in 54 tissue types.

Problems

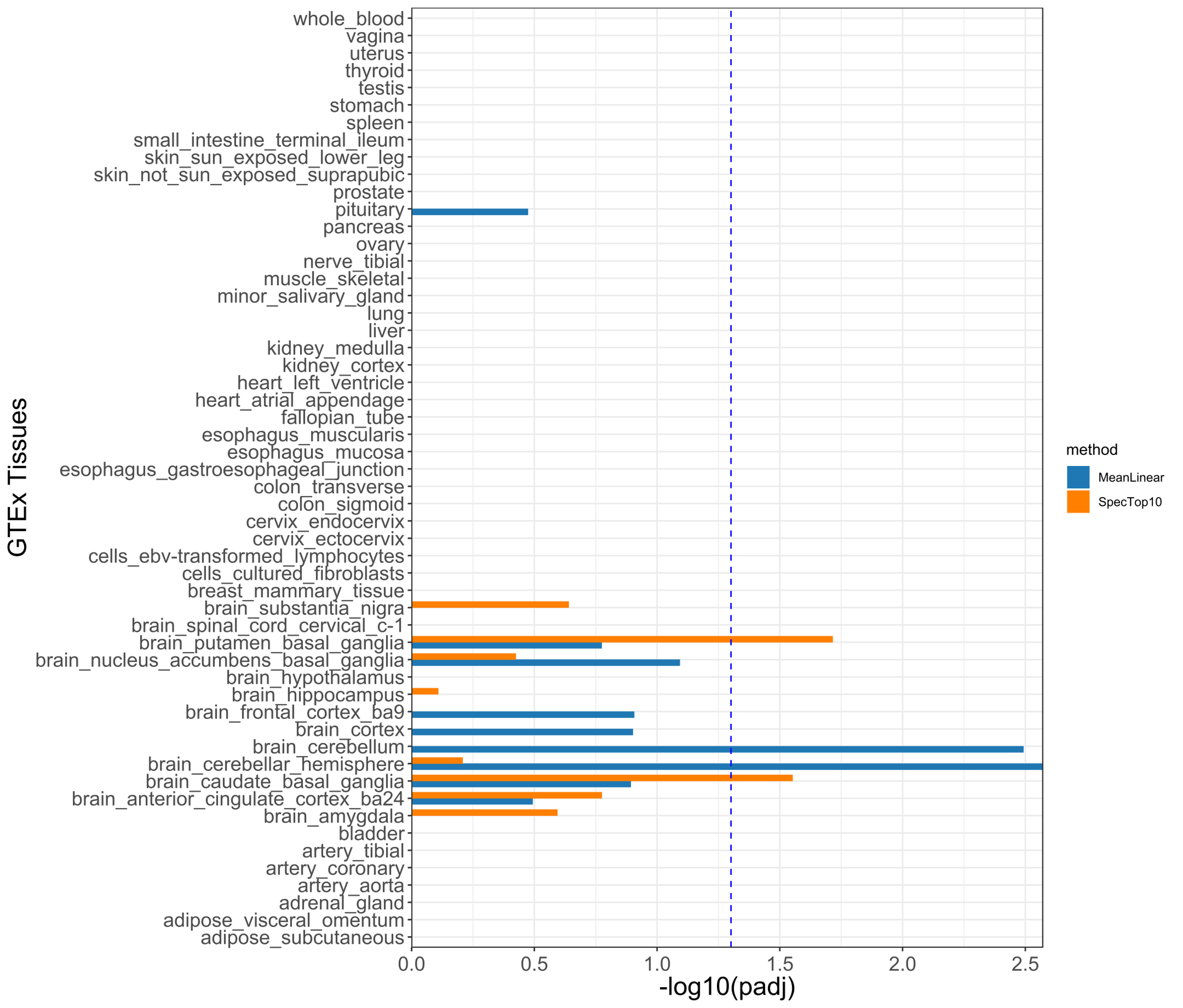

BeerPref

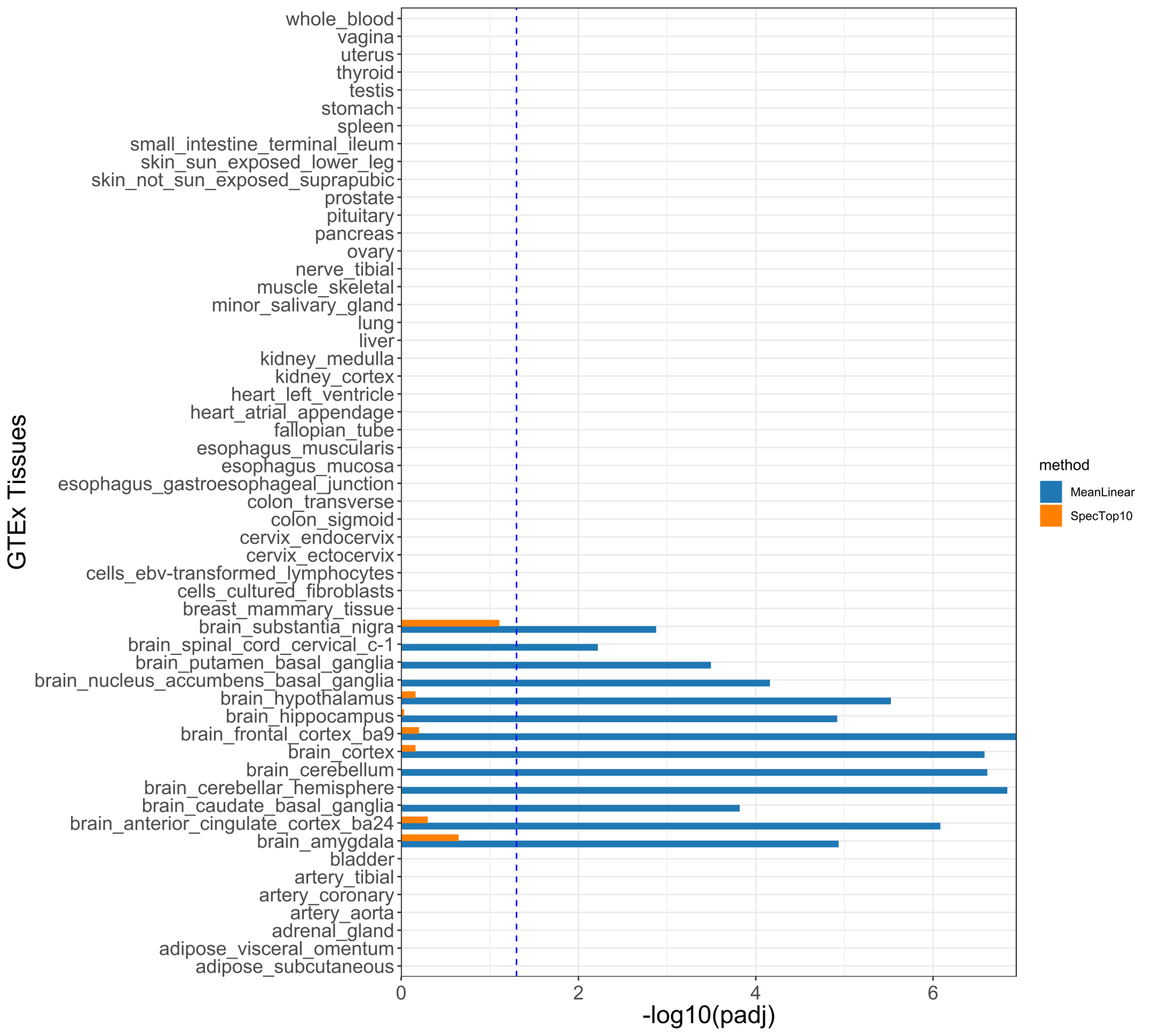

Consumption

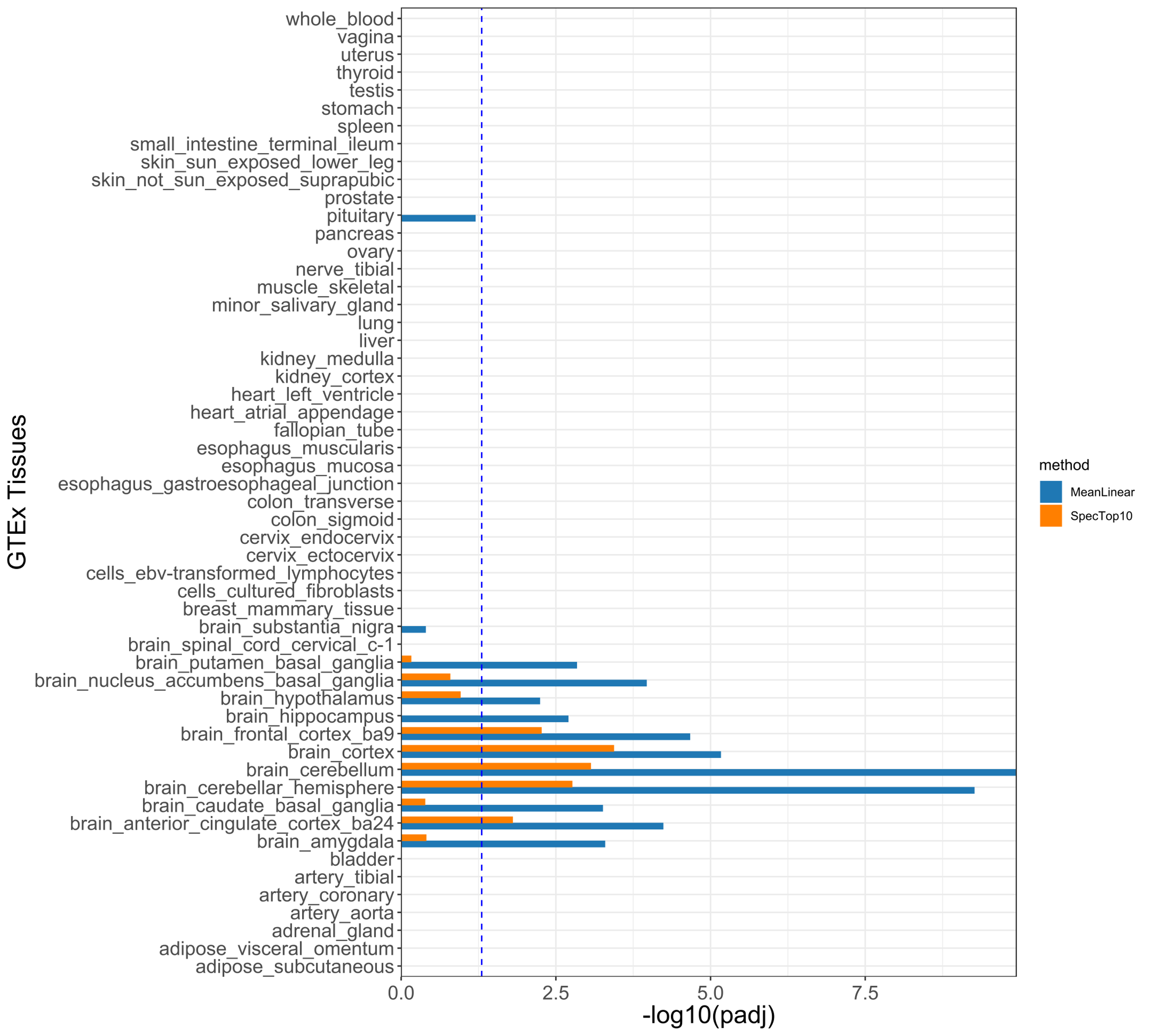

AtypicalPref

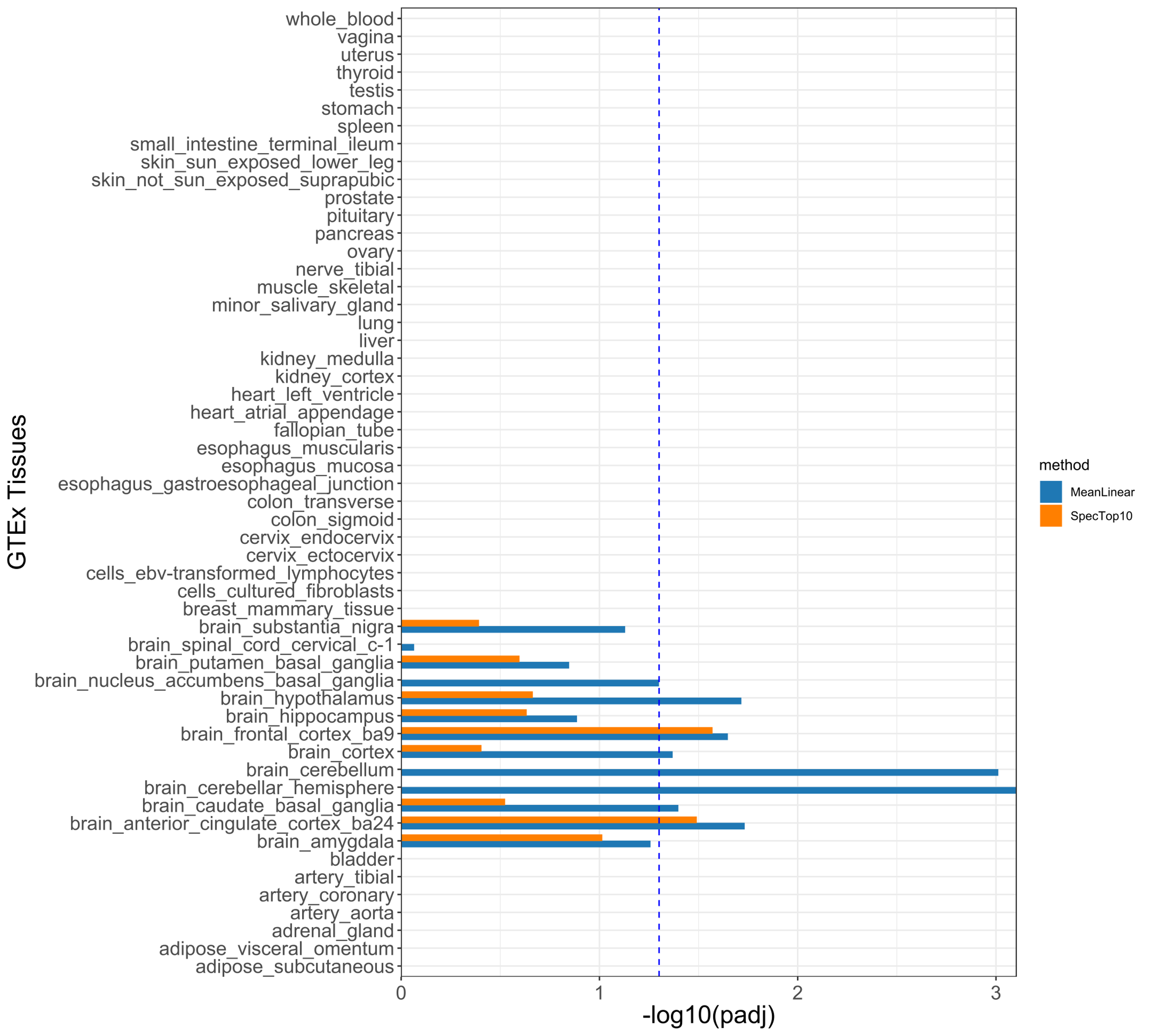

*Note: MeanLinear tests (blue bars) examine whether the gene-based aggregate association signal predicts mean expression of genes in a certain tissue. SpecTop10 tests (orange bars) examine whether gene-based association signal is enriched in genes that are specifically expressed in that same tissue (top 10% of genes with specific expression in target versus other tissues). The blue dashed line represents the significance threshold after Bonferroni correction for 54 tissue types.*

##### Supplementary Figure S6. Enrichment of GWAS association signal for the latent factors in genes with cell type specific expression in 10 regions of the adult human brain.

Problems

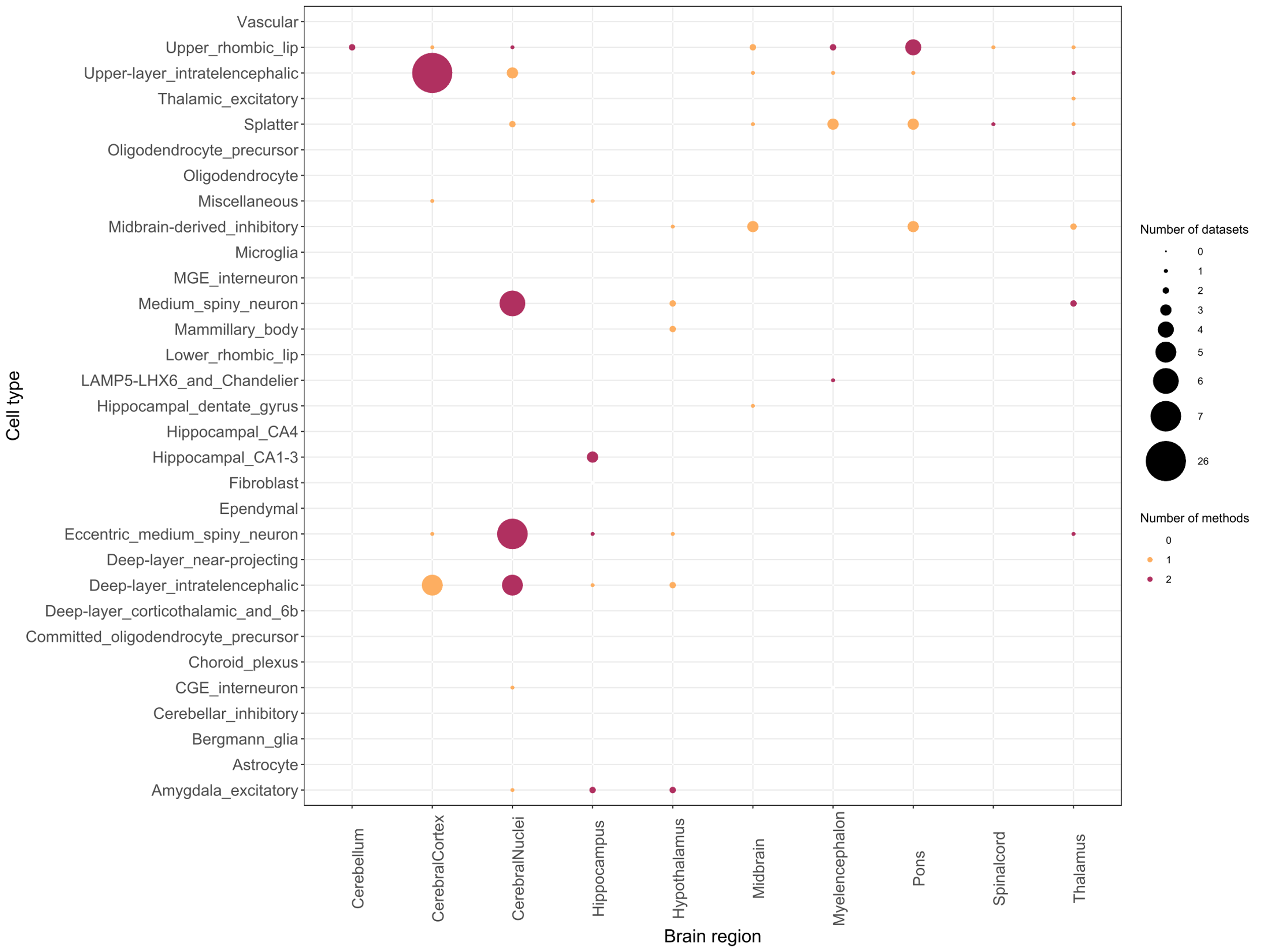

BeerPref

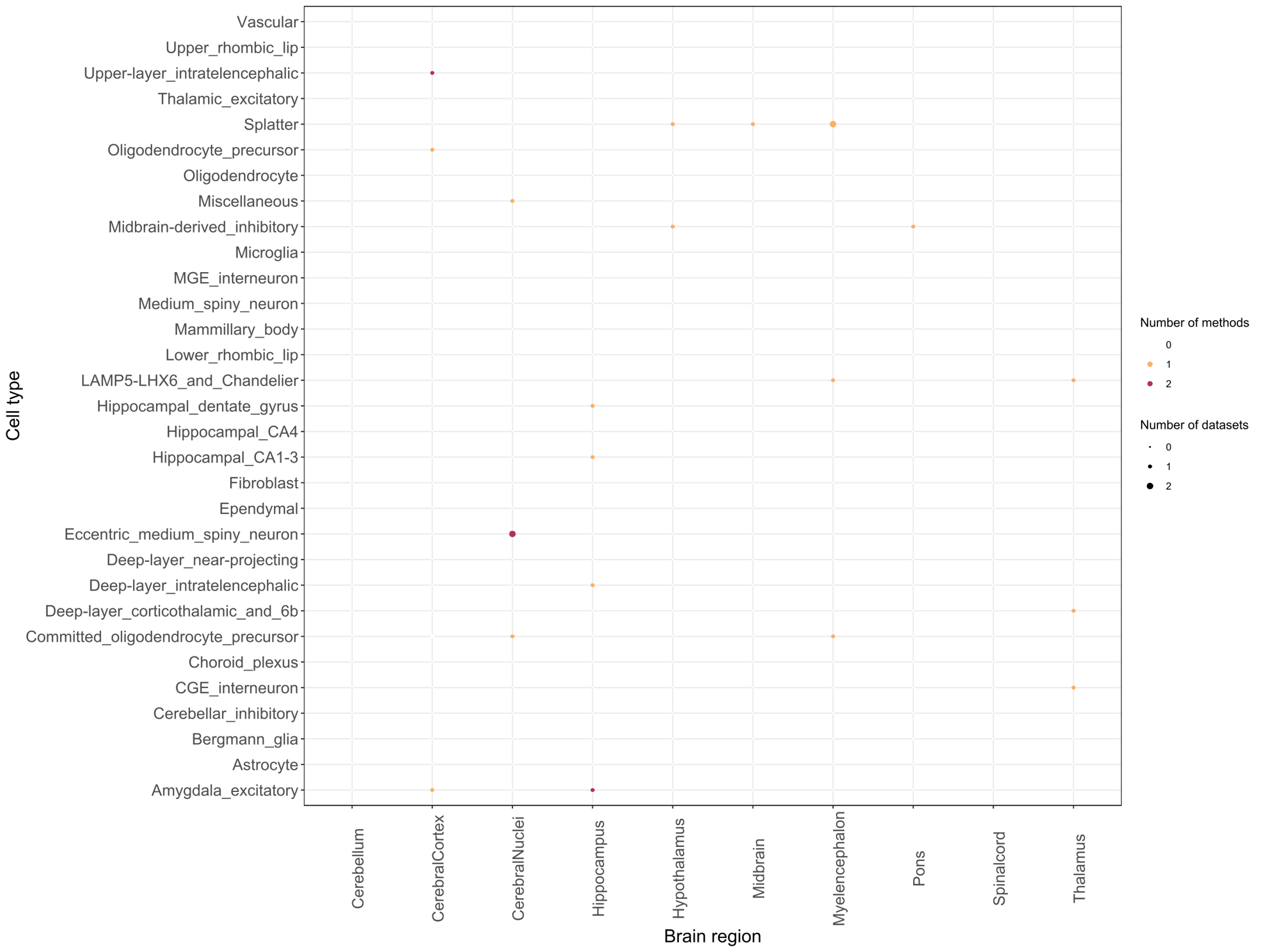

Consumption

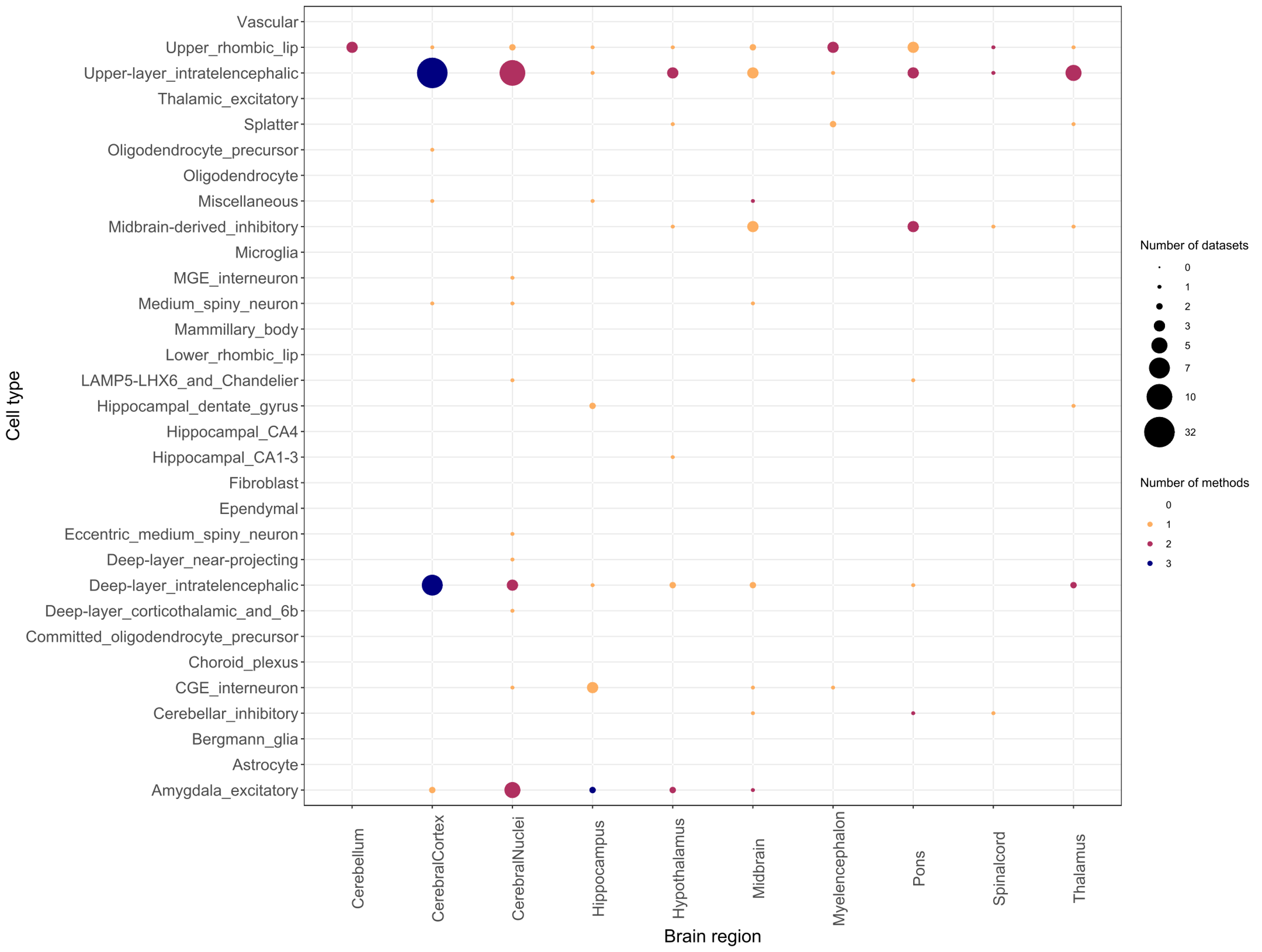

AtypicalPref

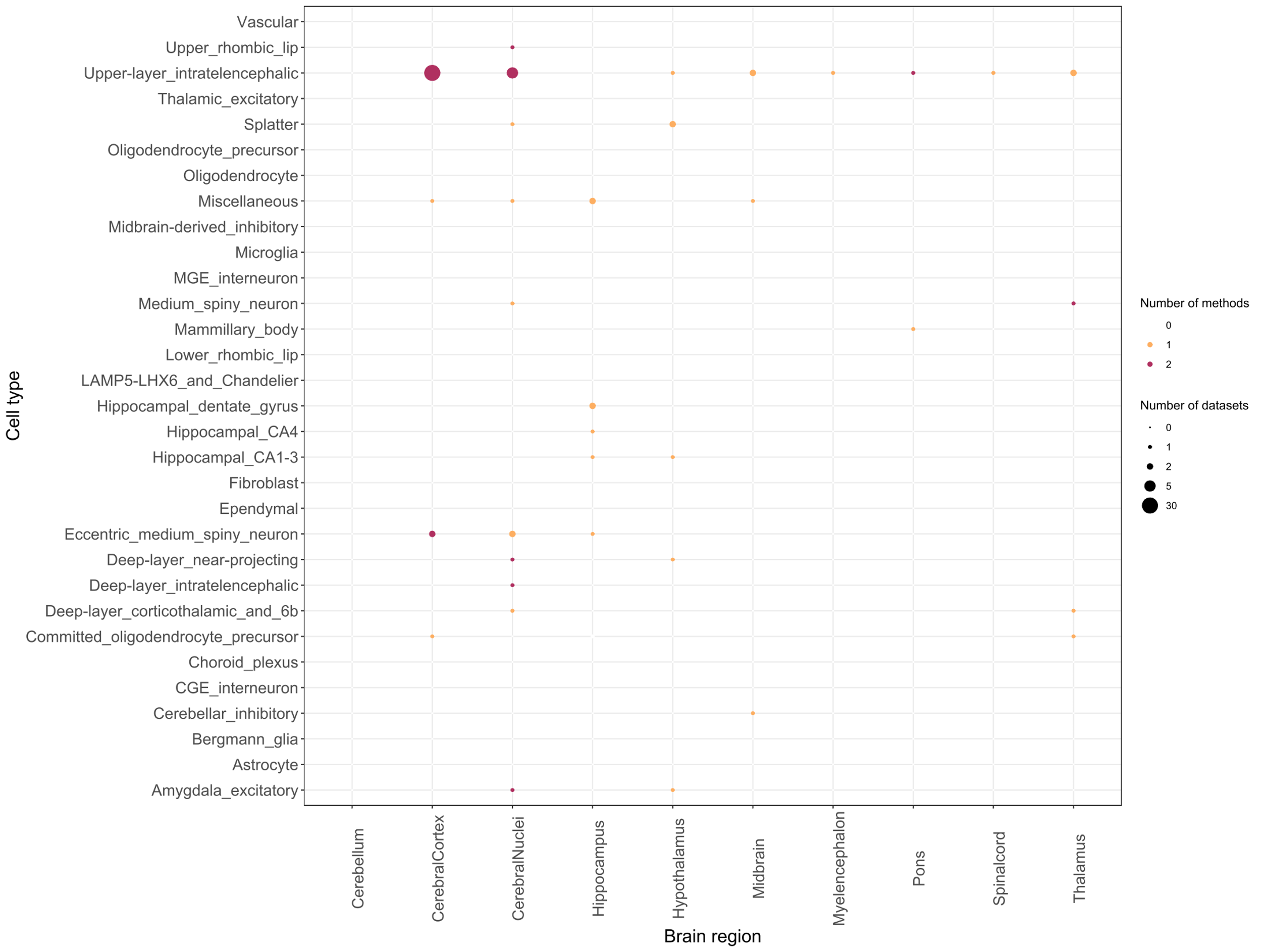

*Note: Multiple dissections were performed within brain regions and each dissection was considered a “dataset” (Siletti et al., 2022). Four methods for testing cell type specificity were carried out (FUMA Celltype, MAGMA Celltyping, CELLECT-MAGMA, and CELLECT-LDSC) and Bonferroni correction was applied for the number of cell types examined within each method within each dataset. Enrichment was considered significant if the test was significant in the same cell type across multiple methods and/or datasets.*

##### Supplementary Figure S7. Phenome-wide association (PheWAS) of polygenic scores from AUB latent factors predicting medical conditions in the BioVU cohort.

a) Problems

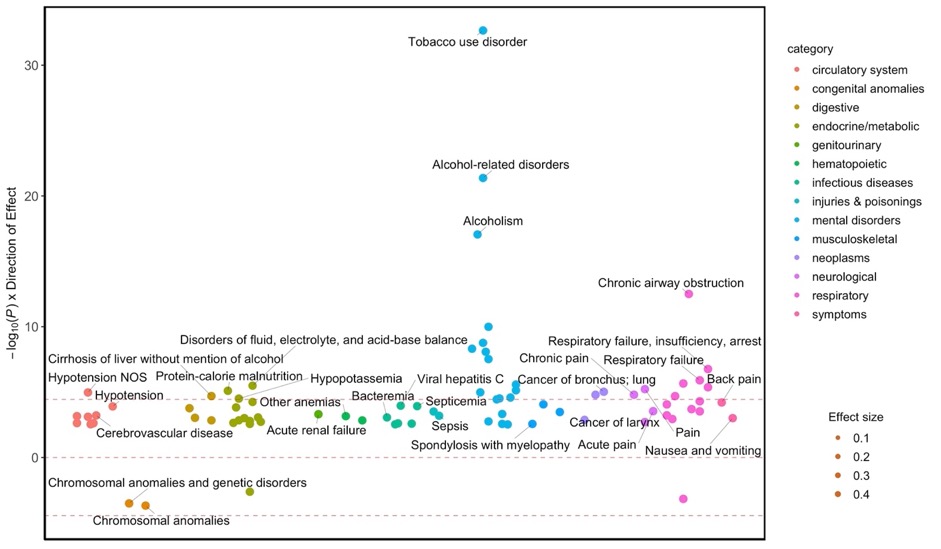

b) BeerPref

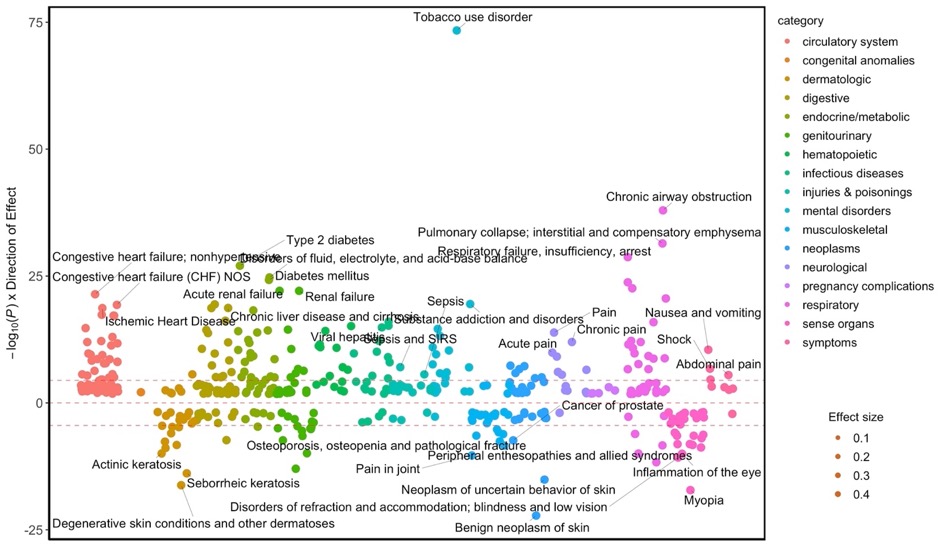

c) Consumption

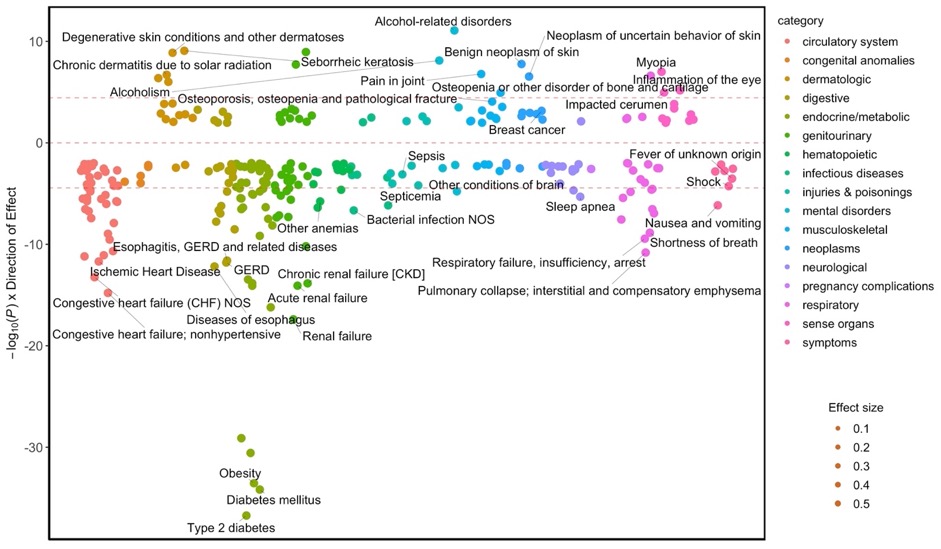

d) AtypicalPref

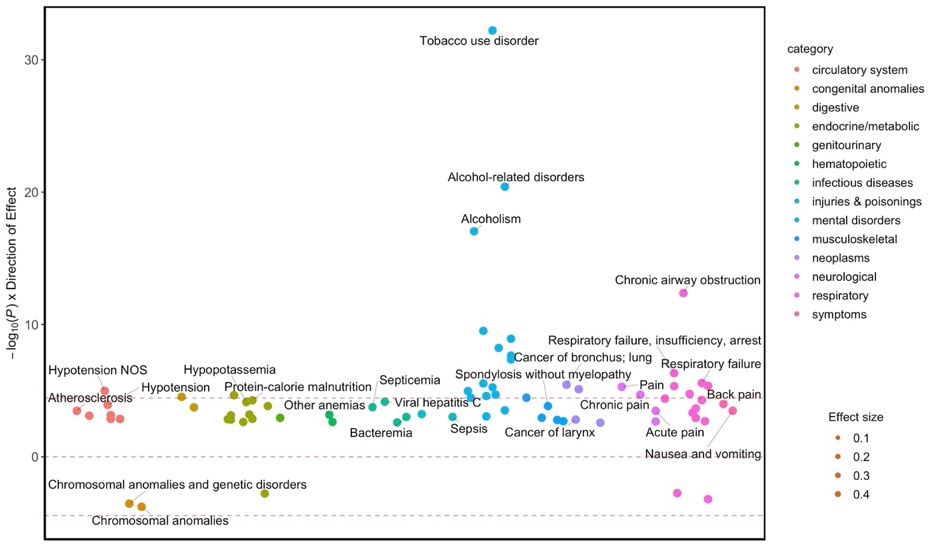

##### Supplementary Figure S8. Phenome-wide association (PheWAS) of polygenic scores from AUB latent factors predicting medical conditions in the MGBB cohort.

a) Problems

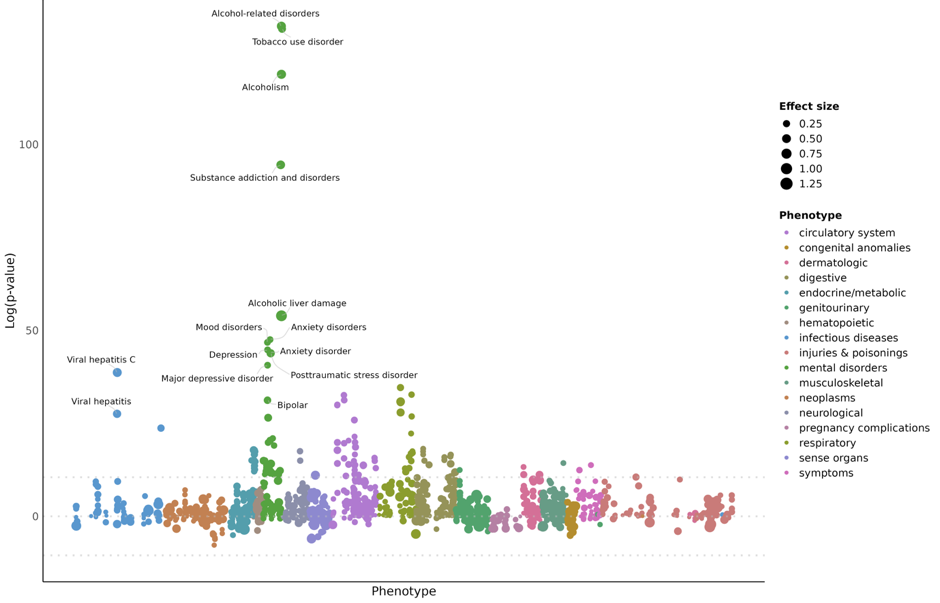

b) BeerPref

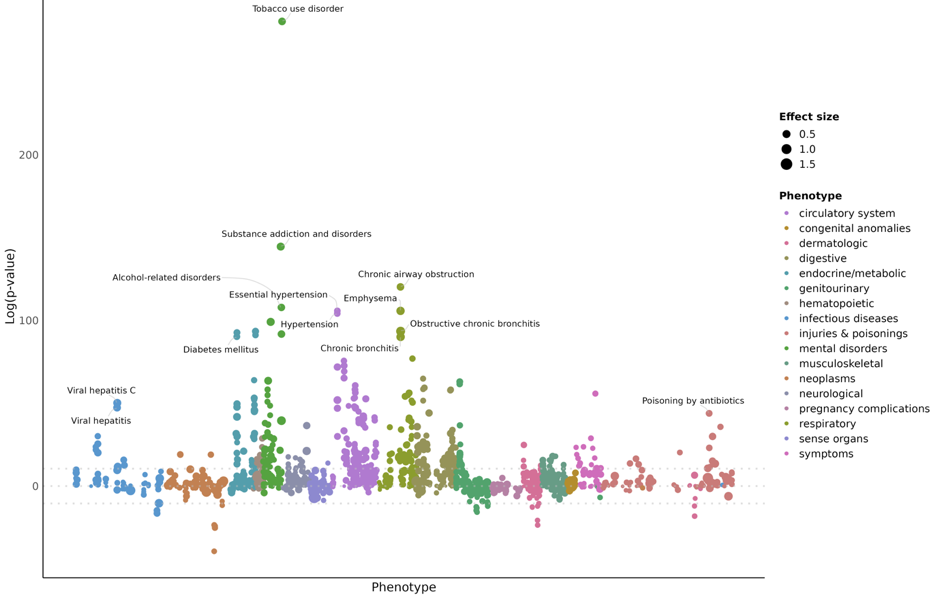

c) Consumption

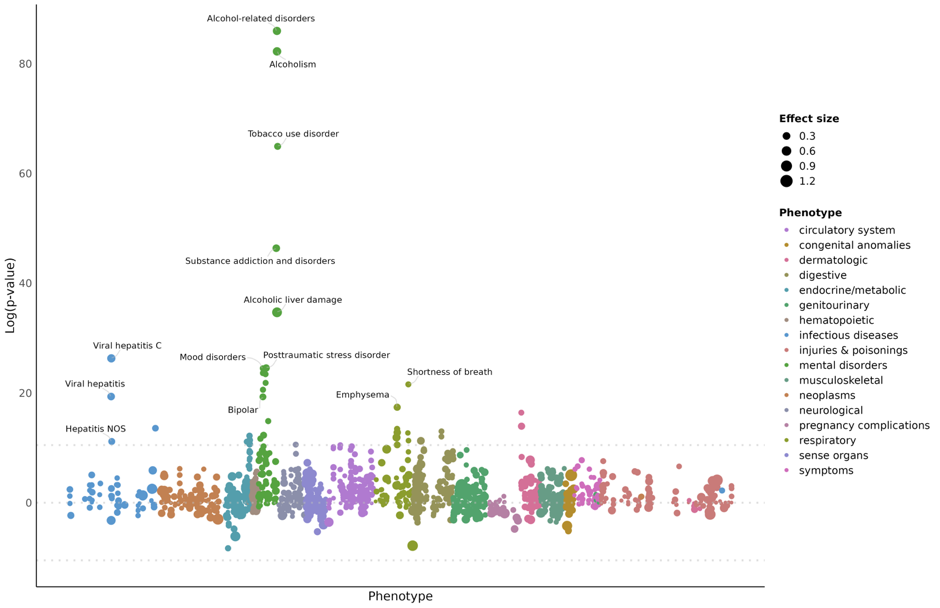

d) AtypicalPref

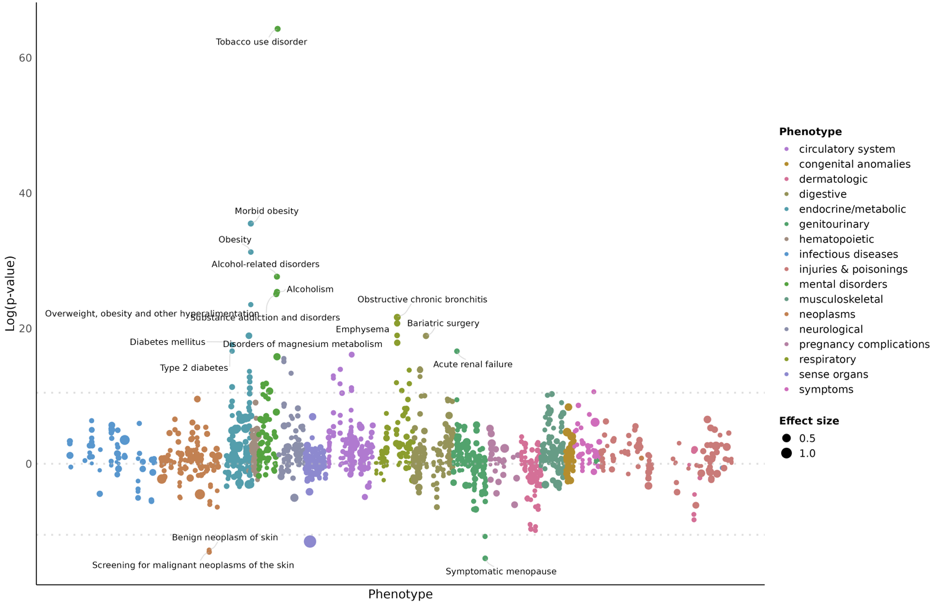
